# Human CSF proteogenomics links genetic variation to neurodegenerative disease proteins

**DOI:** 10.64898/2026.02.12.26345733

**Authors:** Raquel Puerta, Pablo García-González, Itziar de Rojas, Maria Capdevila-Bayo, Clàudia Olivé, Álvaro Muñoz-Morales, Paula Bayón-Buján, Alejandro Valenzuela, Chengran Yang, Jigyasha Timsina, Menghan Liu, Sathyaseelan Chakkarai, Oscar Sotolongo-Grau, Berta Calm, Andrea Miguel, Ariadna Solivar, Laura Montrreal, Marta Martínez, Asif Khan, Feiyang Zhao, Natàlia Tantinyà, Maitée Rosende-Roca, Montserrat Alegret, Sonia Moreno-Grau, Maria Victoria Fernández, Marta Marquié, Sergi Valero, Jose Enrique Cavazos, Pilar Sanz, Xavier Montalban, Lluis Tàrraga, Bart Smets, Mercè Boada, Sudha Seshadri, Muralidharan Sargurupremraj, Carlos Cruchaga, Amanda Cano, Alfredo Cabrera-Socorro, Agustín Ruiz

**Affiliations:** Ace Alzheimer Center Barcelona – Universitat Internacional de Catalunya, Spain; Doctorate in Biotechnology, Facultat de Farmàcia i Ciències de l’Alimentació, Universitat de Barcelona, Avda. Diagonal 643, 08028 Barcelona, Universitat de Barcelona, Spain; CIBERNED, Network Center for Biomedical Research in Neurodegenerative Diseases, National Institute of Health Carlos III, Madrid, Spain; Luxembourg Centre for Systems Biomedicine, University of Luxembourg, Luxemburg; Departament de Farmacologia, Toxicologia i Química Terapèutica, Facultat de Farmàcia i Ciències de l’Alimentació, Universitat de Barcelona, Barcelona, Spain; Department of Psychiatry, Washington University School of Medicine, St. Louis, MO, USA; NeuroGenomics and Informatics Center, Washington University School of Medicine, St. Louis, MO, USA; Glenn Biggs Institute for Alzheimer’s & Neurodegenerative Diseases, University of Texas Health Science Center, San Antonio, TX, USA; Johnson & Johnson, Neuroscience Discovery.; South Texas Alzheimer’s Disease Research Center, San Antonio, TX, USA; Department of Neurology and Multiple Sclerosis Center of Catalonia (Cemcat), Vall d’Hebron University Hospital, Universitat Autònoma de Barcelona (UAB), Barcelona, Spain; National Heart, Lung, and Blood Institute’s Framingham Heart Study; Framingham, Massachusetts 01702, United States; Department of Neurology, Boston University Chobanian & Avedisian School of Medicine; Boston, Massachusetts 02118, United States; Hope Center for Neurological Disorders, Washington University School of Medicine, St. Louis, MO, USA; Charles F. and Joanne Knight Alzheimer Disease Research Center, Washington University School of Medicine, St. Louis, MO, USA; Department of Microbiology, Immunology and Molecular Genetics. Long School of Medicine. University of Texas Health Science Center, San Antonio, Texas, USA

**Author notes:** **Corresponding author**: **Agustín Ruíz**, Ace Alzheimer Center Barcelona. C/ Marquès de Sentmenat, 57, 08029, Barcelona, Spain. Last co-authors have contributed equally.

**Keywords:** proteomics, genomics, SOMAscan, pQTLs, GWAS, cerebrospinal fluid, neurodegeneration

## Abstract

The cerebrospinal fluid (CSF) proteome offers a direct readout of central nervous system (CNS) biology but its genetic architecture remains incompletely defined. We conducted the largest single-site CSF genome-wide association study (GWAS) to date, analysing 7,092 SomaScan proteins in 1,259 individuals. Using a covariate-adjusted model including proteomic PCs and disease status, we identified 1,971 genome-wide significant pQTLs (954 cis, 971 trans), 1,409 of which replicated in an independent CSF dataset. We discovered 264 previously unreported loci, replicated 511 associations, refined 80 known loci, and 265 proxy-based associations. Using a previously published reproducibility framework, we show that robust discovery concentrates in reliable measurements, underscoring the importance of rigorous quality control. Enrichment analyses revealed immune/complement and extracellular matrix biology. Mendelian randomization prioritised causal proteins: PILRA, TREM2, IL34, CR2, SHARPIN and ERBB1 (Alzheimer’s disease); BST1 and GPNMB (Parkinson’s disease); STX6 (Creutzfeldt Jacobs disease); and ATXN3 and B4GALNT1 (Amyotrophic lateral sclerosis), providing a scalable framework for orthogonal target validation in neurodegeneration.

**Highlights:** CSF Aβ42 and p-tau, CSF total protein and Qalb are major contributors to the proteomic variance and may act as potential confounders.

Most pQTLs were found in proteins classified as “reproducible” based on our CSF proteomic score.

The most stringently adjusted GWAS model maximized pQTL discovery among the highest-reproducibility proteins (score 1 and A).

We identified 264 novel CSF pQTLs that were not described in previous analyses, replicated 511 CSF pQTLs, 80 map refinements and 265 proxy SNPs, predominantly involved in immune-related, inflammatory, and extracellular matrix mechanisms.

We found 281 CSF pQTLs that were also systemic modulators of plasma protein levels that were enriched in immune-related and extracellular matrix mechanisms.

We identified and validated several causal proteins associated with AD and other neurodegenerative disorders.

## Main

Alzheimer’s disease (AD) is the most common cause of dementia, creating a massive socioeconomic burden with an expected increase in prevalence over the next 20 years due to aging populations^1–3^. To address this significant health challenge, large efforts have been conducted to understand the molecular mechanisms underlying complex multifactorial diseases and identify novel biomarkers. In this sense, Genome-Wide Association Studies (GWAS) have led to the discovery of multiple single nucleotide polymorphisms (SNP) associated with neurodegenerative disorders, including AD^4–9^. The aetiology of these complex, multifactorial diseases remains only partly resolved. Beyond risk loci, genetic studies have enabled systematic interrogation of disease-relevant endophenotypes, clarifying the genetic architecture of quantitative traits that closely track underlying pathology^10,11^. However, while GWAS have helped to pinpoint genetic risk markers, they are often insufficient on their own to identify the causal variant or elucidate the molecular mechanisms at play. Therefore, proteogenomic approaches, integrating GWAS and high-throughput proteomic profiling, could be of high interest to dissect AD pathological mechanisms and identify potential biomarkers.

Recent advances in multiplexed, high-throughput proteomic technologies enable parallel quantification of thousands of proteins^12^. The dominant platforms include DNA aptamers-based assays (SomaScan), or antibody-based approaches (Olink)^13,14^. To date, most large-scale proteomic studies have focused on plasma or serum due to biomaterial accessibility. By contrast, CSF remains comparatively under-studied despite being a scarce biofluids that provides direct insight into CNS pathological processes^15^. Large consortium efforts, including the Global Neurodegeneration Proteomics Consortium (GNPC), have boosted power through joint analyses of thousands of samples. However, only a subset has matched genotyping, limiting fully integrated proteomic analyses^16,17^.

Integrating genomic and proteomic data has enabled systematic mapping of protein quantitative trait loci (pQTLs), genomic regions modulating protein abundance^18,19^, accelerating proteome characterization and mechanistic insight into disease. pQTLs are tipically classified into cis effects (variants near the gene encoding the measured protein), or trans effect (distal, unlinked loci)^20^. Multiple plasma proteogenomic analysis have been conducted considering several proteomic strategies such as SomaScan 5k and 7k^19–21^. Moreover, we and others have extended their proteogenomic evaluations of the CSF proteome using the proteomic platforms SomaScan 1.1k, 5k and 7k^18,22–24^, multiple Olink proteomic panels^25^, the Human DiscoveryMAP Panel and a Luminex 100 platform^26^. Because of its direct contact with the CNS, the CSF biofluid is remarkably important in the study of AD and other neurodegenerative diseases providing highly valuable information about pathological changes occurring through the disease development^3,27,28^.

Here, we map the CSF proteogenomic landscape in the large ACE Alzheimer Center Barcelona (ACE) cohort, integrating genome-wide genotypes with CSF SOMAscan 7k (v4.1) profiling in 1,370 individuals (>7,000 CSF analytes). We systematically identify loci regulating CSF protein abundance, classify genetic variants as cis- or trans-pQTLs, and investigate these markers as instruments to test causal links to AD, and other neurodegenerative diseases.

## Results

### Proteomic Characterization, PCA Exploration, and Demographics

Following proteomic quality control (QC), we analysed 1,259 well-characterised individuals from the ACE CSF cohort with matched genomic and CSF proteomic information. Consistent with typical AD cohorts, women were over-represented (57.7%), and most participants had mild cognitive impairment (MCI). The mean age was of 72.40 ± 8.84 years and the mean Mini-Mental State Examination (MMSE) was 24.40 ± 4.44. As expected, we observed lower MMSE values in dementia cases compared with healthy controls (HC) and MCI individuals. All participants were assigned an AT(N) classification based on CSF biomakers^29^. The cohort was enriched for A-T-N- (n=29.80%) and A+T+N+ (n=33.30%) profiles. Additionally, 34.60% carried at least one *APOE* ε4 allele (**Supplementary Table 1**).

We then subjected SomaScan CSF measurements to a stringent QC and reproducibility filtering as previously described^30^. This framework stratifies proteins by measurement reproducibility and reliability (Methods, **Supplementary Table 2**). After excluding non-human and low-confidence measurements, we retained 7,092 high-confidence proteins for downstream analysis (**Figure 1A-B**).

**Figure 1.**
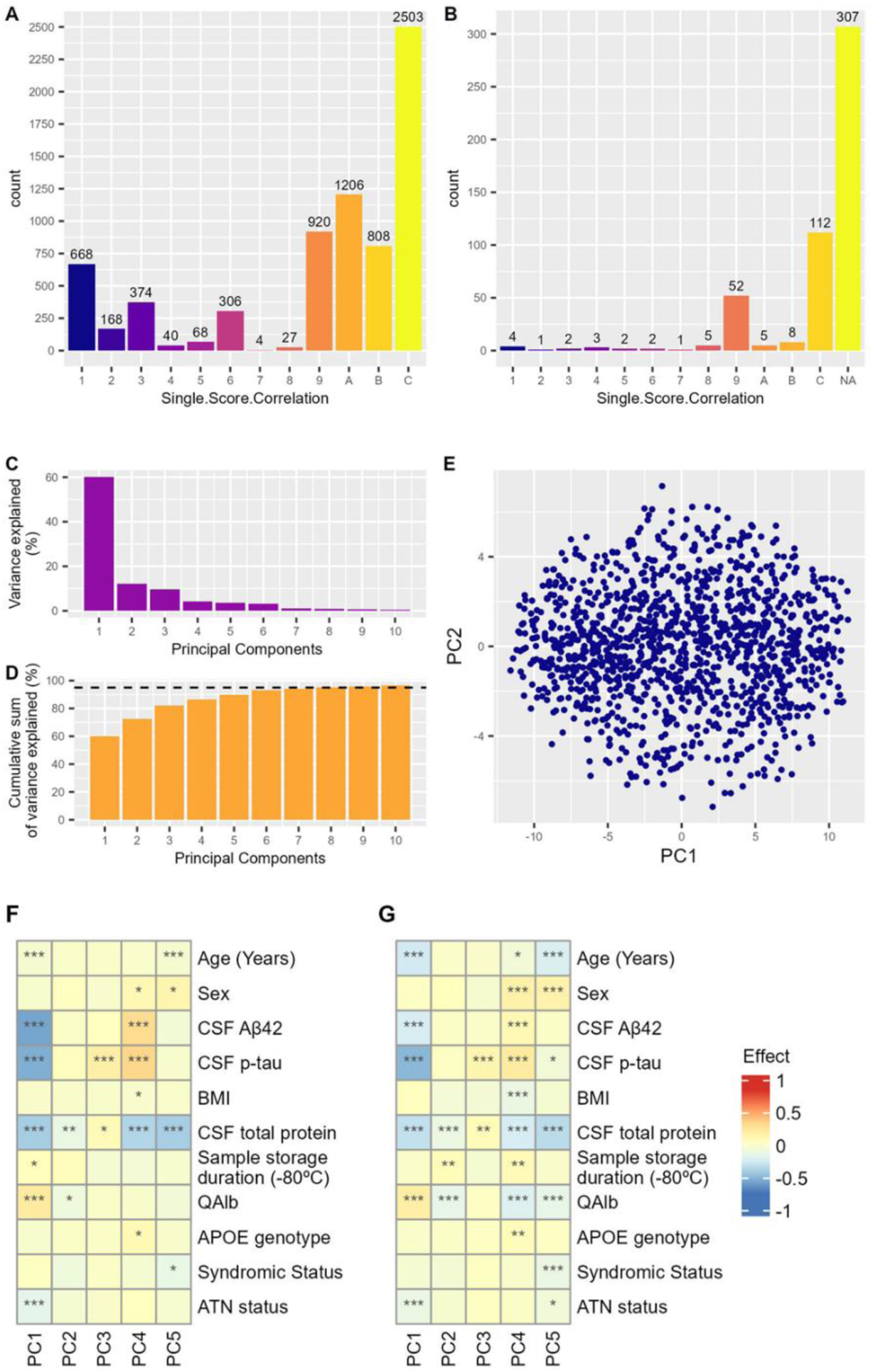
Summary of CSF SomaScan proteomic QC, and PCs characterization. A) Summary of the 7,092 proteins that passed the QC. B) Summary of the 504 proteins that failed the QC by Puerta and Cano et al. Reproducibility Score. Note that non-human proteins were not included in the reproducibility score calculation (NA)^30^. C) Variance explained by the top 15 PCs. D) Cumulative sum of variance explained by the top 15 PCs. E) Graphical representation of each individual PC1 and PC2. F) Linear regression analysis of the first 5 PCs adjusted by all phenotypes. G) Pearson correlation of the first 5 PCs as a non-adjusted model. An asterisk (*) represents a p value lower than 0.05, two asterisks (**) represent a p value lower than 0.01, and three asterisks (***) represent a p value lower than 0.001.

To account for major sources of confounding in the CSF proteomic data, we performed Principal Component Analysis (PCA) and investigated which clinical, demographic, and AD biomarker variables were associated with principal components (PCs). The first two components (PC1 and PC2) dominated the structure of the data, explaining 60.17% and 12.19% of total variance, respectively (**Figure 1C-E**). Variation along these axes was driven primarily by barrier function and AD pathology, with the strongest associations observed for CSF Aβ42, CSF p-tau, total CSF protein, and the albumin index (Qalb). Conversely, age, sex, and clinical diagnosis showed no clear associations, whereas proteomic PCs were closely aligned with AT(N) profile (**Figure 1F-G, Supplementary Figure 1,** Methods).

### Genetic Architecture of the CSF Proteome

Across 21,276 protein-wise GWAS (7,092 proteins × 3 adjustment models), we detected ∼2,000 genome-wide significant pQTLs: 1,879 (811 cis-pQTLs, 1,028 trans-pQTLs), 1,998 (947 cis-pQTLs, 1,011 trans-pQTLs) and 1,971 (954 cis-pQTLs, 971 trans-pQTLs) for models 1-3, respectively (Methods, **Supplementary Table 3-5**). While comparing the findings of the three models, we observed a substantial overlap between pQTL-protein pairs in models 2 and 3 (52.55%), suggesting that the inclusion of covariates such as age, sex, disease status and genomic PCs have a modest impact on our results compared to the proteomic PCs. The majority of aptamers were found by the 3 models (63.73%), with a greater overlap in aptamers identified by models 2 and 3 (77.82%) (**Figure 2A-B**). Considering reproducible proteins^30^, similar results were found with a greater overlap of pQTL-protein pairs (51.05%) and aptamers represented (81.70%) between models 2 and 3 (**Figure 2C-D**).

**Figure 2.**
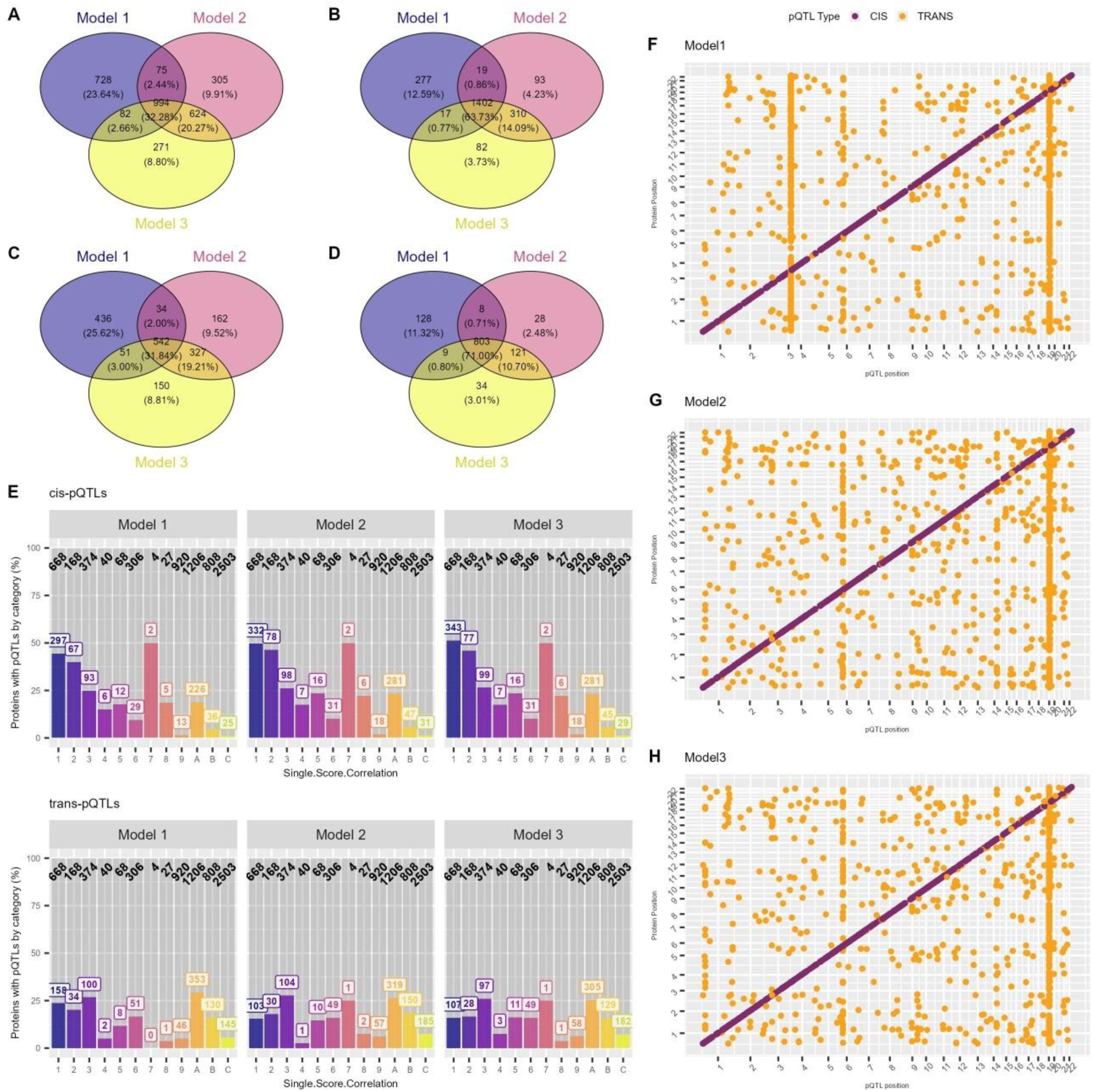
Representation of the CSF pQTL results across GWAS models. A) Venn diagram comparing the unique protein-pQTLs pairs. B) Venn diagram comparing the unique aptamers with pQTLs. C) Venn diagram comparing the unique protein-pQTLs pairs considering proteins with the highest reproducibility proteins. D) Venn diagram comparing the unique aptamers with pQTLs considering proteins with the highest reproducibility proteins. E) Percentage of proteins with cis- and trans-pQTLs classified by Single.Score.Correlation and model. The black labels refer to the total number of proteins included in each reproducibility or Single.Score.Correlation category^30^. F) pQTL map (model 1). G) pQTL map (model 2). H) pQTL map (model 3). The genome wide-significance threshold was set to 5e-08 for cis-pQTLs and 6.25e-09 for trans-pQTLs. Cis-pQTLs are coloured in purple and trans-pQTLs in orange.

Using our previously described reproducibility framework^30^, reproducible proteins yielded more pQTLs than lower-confidence categories, with the strongest signal under the most stringently adjusted model (model 3). In this sense, the highest-reproducibility proteins (score 1 and A^30^) were enriched for cis-pQTLs, and showed a relative depletion of trans-pQTLs (n_CIS_=624; n_TRANS_=412; **Supplementary Table 6,** **Figure 2E**). Cis-pQTLs were preferentially observed among reproducible proteins that also correlate with antibody-based measurements (Olink), whereas trans-pQTLs were enriched among reproducible proteins without cross-platform concordance (**Supplementary Table 6**).

In GWAS model 1, we detected three prominent trans-pQTL regions on chromosomes 3 (near *GMNC*), 6 (*HLA*), and 19 (*APOE*) (**Figure 2F**). Interestingly, in models 2 and 3, which include the proteomic PCs covariates, only the chromosomes 6 and 19 trans-signals remained (**Figure 2G-H**). Together, these patterns suggest that a subset of trans associations, particularly those enriched in proteins lacking cross-platform concordance, may reflect platform-specific technical effects such as reagent- or assay-dependent biases rather than true distal genetic regulation. Consistent with this interpretation, the inclusion of proteomic PC covariates attenuated several prominent trans-signals (**Figure 2F-H**), indicating that latent proteomic structure can absorb broad non-biological variation.

We considered the model 3 pQTL results for further analysis. The χ^2^ test revealed that proteins in highly reproducible categories (1/A) were significantly more likely to have pQTLs than those in non-reproducible categories (9/C) (1-9 P=0.004, 1-C P=9.30e-05, A-9 P=0.007, A-C P=1.34e-04), whereas no significant differences were observed within reproducible strata (1-A P=0.76, 9-C P=1.00). These results support the validity of our reproducibility score for prioritizing reliable proteins measurements in CSF proteogenomic analyses (**Supplementary Table 7**).

### pQTL identification and independent replications

To systematically explore replications of our CSF pQTLs, we considered the Washington University School of Medicine (WashU) dataset removing any overlap with our study (Methods). Out of the 1,971, 1,409 pQTLs (71.49%) were found in the independent replication at P<0.05 (WashU, **Supplementary Figure 2**). Among these, we identified 264 (98 cis-, 161 trans-pQTLs) novel pQTLs, representing completely new associations not previously reported in the literature. Notably, 161 of them were not described in the largest CSF pQTLs study to date^24^, and 103 were in low linkage disequilibrium (LD) (R2<0.5) with their reported pQTL. This suggest that the addition of extra covariates, such as proteomic PCs, might lead a refined view of this genomic regions (**Figure 4C, Supplementary Table 8**). We also replicated 511 pQTLs (237 cis-, 267 trans-pQTLs) where the same lead SNP at a previously reported locus^24^ was successfully replicated across our analyses. High concordance was observed in the betas (effect size estimate) between the ACE and the WashU replication (Pearson R=0.98) (**Figure 4A-B**). These results confirm the involvement of previously reported loci in regulating CSF protein levels, and support the robustness of SNP-protein associations across studies. Overall, replication was more frequent for cis-pQTLs than for trans-pQTLs, with one notable exception: a strong trans-signal at chromosome 19 (**Figure 4D**). In addition, 265 pQTLs (178 cis-, 84 trans-pQTLs) replicated via a proxy SNP in strong LD with the originally reported variant^24^, and 80 pQTLs (53 cis-, 26 trans-pQTLs) reflected fine-mapping/refinement at known loci, where our data identified a more informative marker at a previously known locus (**Figure 4E-F**).

**Figure 3.**
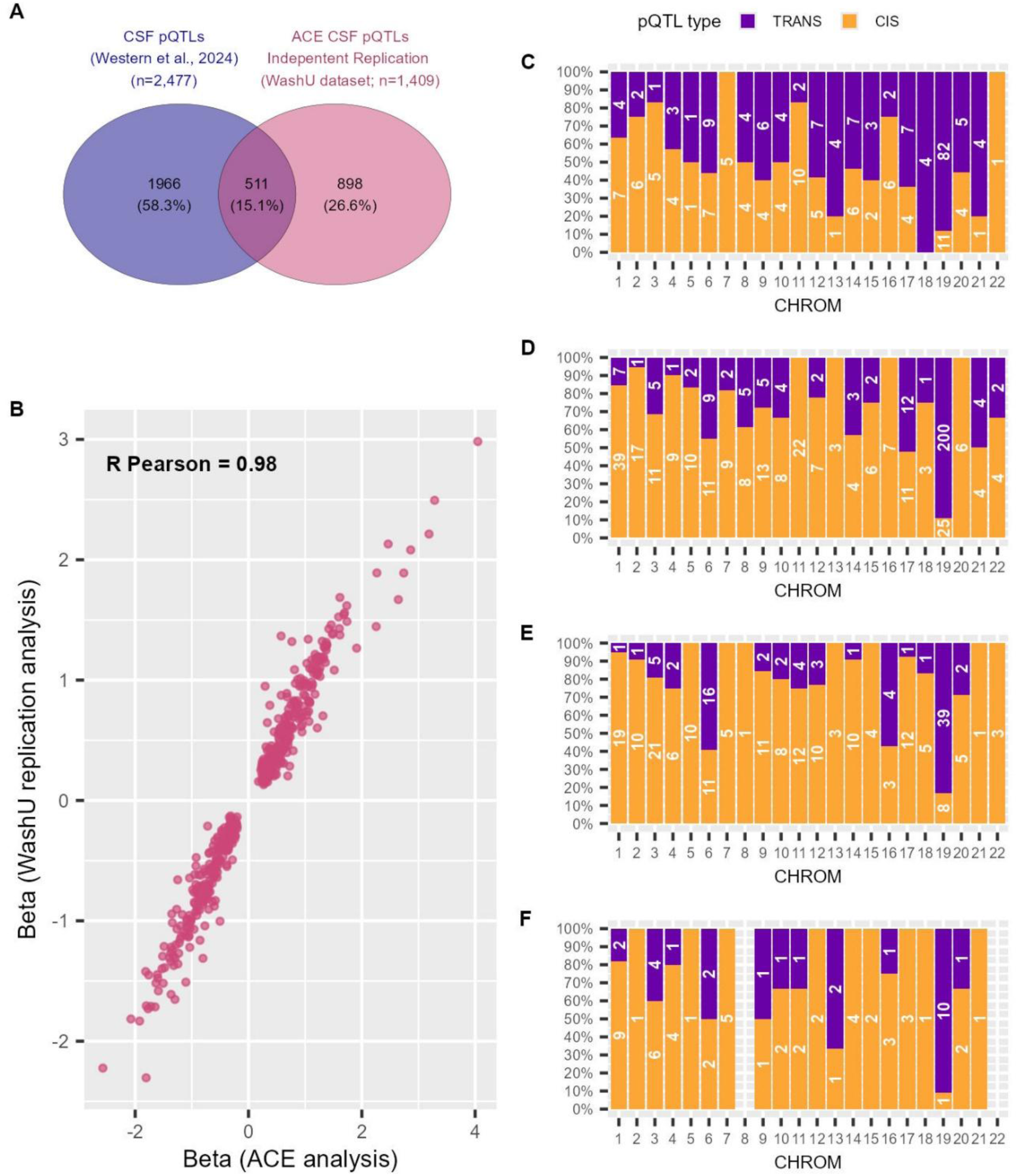
Overview of the replications of the ACE pQTL candidates. A) Venn diagram of genome-wide significant pQTL-protein pairs between ACE independent replication pQTLs (n=1,409; pink), and CSF pQTLs reported by Western et al. (n=2,477; purple). B) Comparison of the beta considering replicated pQTLs between the ACE analysis and the independent replication (WashU, n=511). Pearson correlation was conducted. C) Distribution of the novel pQTLs coloured by the pQTL type (n=264). D) Distribution of the novel pQTLs coloured by the pQTL type (n=511). E) Distribution of the proxy pQTLs coloured by the pQTL type (n = 265). F) Distribution of the map refinement pQTLs coloured by the pQTL type (n = 80). cis-pQTL in orange and trans-pQTL in dark purple.

**Figure 4.**
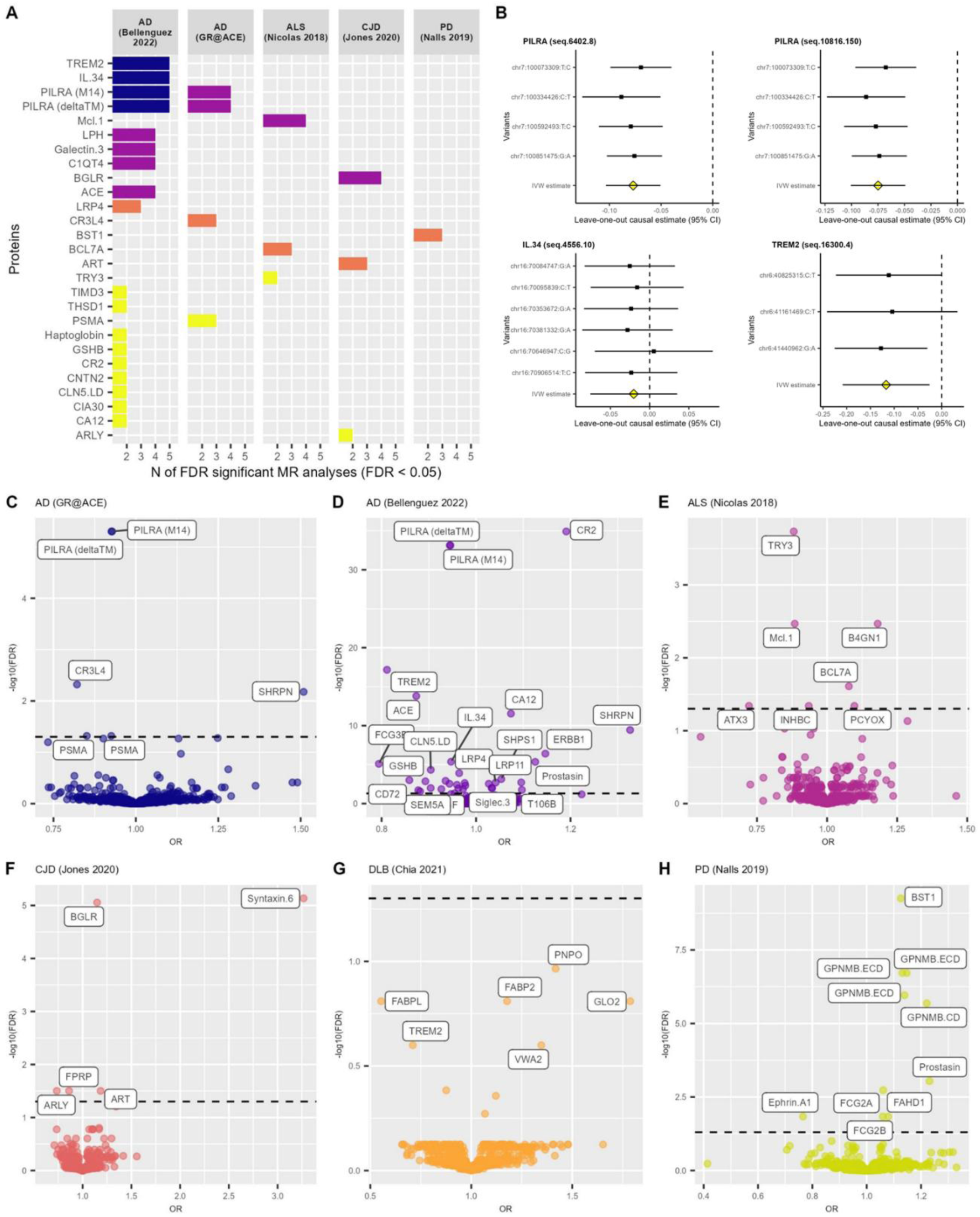
**FDR-significant proteins in MR analyses**. A) Proteins significant in several MR analyses by disease, coloured by the number FDR-significant analyses (FDR<0.05, blue n=5, purple n=4, orange n=3, yellow n=2). B) Leave One Out MR results for 4 proteins FDR-significant in AD in 5 different methodologies (Bellenguez et al.). C) IVW/Wald ratio MR results on AD (GR@ACE). D) IVW/Wald ratio MR results on AD (Bellenguez et al.). E) IVW/Wald ratio MR results on ALS (Nicolas et al.). F) IVW/Wald ratio MR results on CJD (Jones et al.). G) IVW/Wald ratio MR results on DLB (Chia et al.). H) IVW/Wald ratio MR results on PD (Nalls et al.). The significance threshold was set to FDR<0.05 (dashed line).

The remaining 851 signals were classified as “NA” for replication: 562 associations were tested in the independent CSF dataset but did not reach significance, and 289 could not be assessed due to missing data despite being significant in the independent CSF replication. Among these replication categories, we observed a comparable proportion of novel pQTLs across the SomaScan reproducibility score^30^ (ranging from 16.67 to 27.83%). Moreover, irrespective of whether pQTLs were cis or trans, the minor allele frequency (MAF) distributions were similar across replication categories (**Supplementary Figure 3**).

#### Biological significance of enrichment mechanisms of pQTLs

To place pQTLs in biological context, we performed pathway enrichment analysis stratified by replication category and pQTL type. Across pQTL mapping to reproducible proteins, signals consistently converged on immune and extracellular matrix (ECM) biology (**Supplementary Figure 4**).

Novel trans-pQTLs were enriched in complement and ECM-linked mechanisms including *Decreased circulating complement factor B concentration* (Enrichment Ratio=267.81, FDR=0.036), *Scavenging by Class A Receptors* (Enrichment Ratio=105.71, FDR=0.018), and *Proteoglycan binding* (Enrichment Ratio=55.79, FDR=0.036). (**Supplementary Figure 4A-B, Supplementary Table 9-13**). In parallel, replicated cis-pQTLs were enriched in immune/ECM annotations, exemplified by *Immunoglobulin binding* (Enrichment Ratio=30.58, FDR=6.69e-04), and *Complement system* (Enrichment Ratio=12.87, FDR= 3.51e-04). Replicated trans-pQTLs additionally highlighted receptor-mediated mechanisms such as *Scavenging by Class A Receptors* (Enrichment Ratio=85.16, FDR=2.34e-02), alongside ECM biology (**Supplementary Figure 4C-D, Supplementary Table 14-18**).

Among replication-by-proxy and map refinements, proxy cis-pQTLs were enriched in ECM/metabolic signatures (*Proteoglycan binding*: Enrichment Ratio=26.97, FDR=0.033) (**Supplementary Figure 4E-F, Supplementary Table 19-23**), while cis map refinements were linked to structural ECM biology (*Molecules associated with elastic fibres*: Enrichment Ratio=71.56, FDR= 3.87e-02) (**Supplementary Figure 4G-H, Supplementary Table 24-28**). Notably, *Collagen-containing extracellular matrix* emerged as recurrent FDR-significant theme across cis map refinements, novel trans-pQTLs, and both replicated cis- and trans-pQTL categories (**Supplementary Figure 6, Supplementary Table 9-28**), and the same overall pattern held when analysing all proteins with pQTLs (**Supplementary Figure 5, Supplementary Table 9-28**).

#### Plasma Replication of CSF pQTLs

A substantial fraction of CSF pQTLs replicated in plasma across two independent cis-pQTL resources: 438 (58.01%) in replication A (WashU) and 421 (81.59%) in replication B (Pietzner *et al.*)^20,24,31^. Plasma and CSF betas were highly concordant (Pearson R_replicationA_= 0.64; Pearson R_replicationB_=0.81). Of these, 281 plasma cis-pQTLs were shared among plasma studies suggesting that are systemic modulators of protein levels (**Supplementary Figure 7, Supplementary Table 29-30**). These pQTLs were enriched in KEGG annotations associated with *Cytokine-cytokine receptor interaction* (FDR_repl.A_=5.53e-07; FDR_repl.B_=5.54e-08), with additional annotations involved in *Complement and coagulation cascades*, *Lysosome*, *Hematopoietic cell lineage*, *Glutathione metabolism*, *Glycosaminoglycan degradation*, *Other glycan degradation* and *Ecm receptor interaction*. In contrast, 317 (41.99%), and 95 (18.41%) pQTLs were not significant in plasma replications A and B, respectively. 76 of these were shared among plasma replications, suggesting that are exclusive of the CSF reflecting specific regulation occurring in the brain. These CNS-exclusive pQTLs are enriched in KEGG pathways associated with *Cytokine-cytokine receptor interaction* (FDR_repl.A_=3.92e-07; FDR_repl.B_=7.44e-05), and *Axon guidance* (FDR_repl.A_=2.10e-03; FDR_repl.B_=3.34e-02) (**Supplementary Figure 7, Supplementary Table 31**).

### Mendelian Randomization Analyses

Having established a high confidence CSF resource, we investigated these variants as genetic instruments to test causal mendelian randomization (MR) hypotheses, and prioritize potential drug targets for neurodegenerative diseases. To explore causal associations of these CSF protein candidates, we conducted several MR analyses considering 1,376 independent cis-pQTLs not mapped to known pleiotropic regions with sufficient strength (F-statistic>10) (Methods). We observed that several proteins were found FDR-significant (FDR<0.05) across multiple methods (**Figure 5A)**, such as the Paired immunoglobulin-like type 2 receptor alpha (PILRA)-isoforms, triggering receptor expressed on myeloid cells 2 (TREM2), and interleukin 34 (IL34) proteins were associated with an AD (Bellenguez)^9^ using 5 MR strategies. Interestingly, the both PILRA isoforms were related with AD in Bellenguez and GR@ACE summary statistics using several methodologies (**Figure 5A-B**, **Supplementary Table 32**).

**Figure 5.** Summary of CSF SomaScan proteomic QC, and PCs characterization. A) Summary of the 7,092 proteins that passed the QC. B) Summary of the 504 proteins that failed the QC by Puerta and Cano et al. Reproducibility Score. Note that non-human proteins were not included in the reproducibility score calculation (NA)^30^. C) Variance explained by the top 15 PCs. D) Cumulative sum of variance explained by the top 15 PCs. E) Graphical representation of each individual PC1 and PC2. F) Linear regression analysis of the first 5 PCs adjusted by all phenotypes. G) Pearson correlation of the first 5 PCs as a non-adjusted model. An asterisk (*) represents a p value lower than 0.05, two asterisks (**) represent a p value lower than 0.01, and three asterisks (***) represent a p value lower than 0.001.

**Figure 6.** Representation of the CSF pQTL results across GWAS models. A) Venn diagram comparing the unique protein-pQTLs pairs. B) Venn diagram comparing the unique aptamers with pQTLs. C) Venn diagram comparing the unique protein-pQTLs pairs considering proteins with the highest reproducibility proteins. D) Venn diagram comparing the unique aptamers with pQTLs considering proteins with the highest reproducibility proteins. E) Percentage of proteins with cis- and trans-pQTLs classified by Single.Score.Correlation and model. The black labels refer to the total number of proteins included in each reproducibility or Single.Score.Correlation category^30^. F) pQTL map (model 1). G) pQTL map (model 2). H) pQTL map (model 3). The genome wide-significance threshold was set to 5e-08 for cis-pQTLs and 6.25e-09 for trans-pQTLs. Cis-pQTLs are coloured in purple and trans-pQTLs in orange.

**Figure 7.** Overview of the replications of the ACE pQTL candidates. A) Venn diagram of genome-wide significant pQTL-protein pairs between ACE independent replication pQTLs (n=1,409; pink), and CSF pQTLs reported by Western et al. (n=2,477; purple). B) Comparison of the beta considering replicated pQTLs between the ACE analysis and the independent replication (WashU, n=511). Pearson correlation was conducted. C) Distribution of the novel pQTLs coloured by the pQTL type (n=264). D) Distribution of the novel pQTLs coloured by the pQTL type (n=511). E) Distribution of the proxy pQTLs coloured by the pQTL type (n = 265). F) Distribution of the map refinement pQTLs coloured by the pQTL type (n = 80). cis-pQTL in orange and trans-pQTL in dark purple.

**Figure 8.** **FDR-significant proteins in MR analyses**. A) Proteins significant in several MR analyses by disease, coloured by the number FDR-significant analyses (FDR<0.05, blue n=5, purple n=4, orange n=3, yellow n=2). B) Leave One Out MR results for 4 proteins FDR-significant in AD in 5 different methodologies (Bellenguez et al.). C) IVW/Wald ratio MR results on AD (GR@ACE). D) IVW/Wald ratio MR results on AD (Bellenguez et al.). E) IVW/Wald ratio MR results on ALS (Nicolas et al.). F) IVW/Wald ratio MR results on CJD (Jones et al.). G) IVW/Wald ratio MR results on DLB (Chia et al.). H) IVW/Wald ratio MR results on PD (Nalls et al.). The significance threshold was set to FDR<0.05 (dashed line).

To maximize coverage across the CSF proteome, we prioritized the IVW/Wald ratio estimates for downstream interpretation. Overall, we found a 34, 6, 7, 5, and 10 FDR-significant proteins associated with AD (Bellenguez), AD (GR@ACE), Amyotrophic lateral sclerosis (ALS), Creutzfeldt Jacobs disease (CJD) and Parkinson’s disease (PD) respectively, with no significant hits for Dementia with Lewy-bodies (DLB) (**Figure 5C-H, Supplementary Table 33**). Two PILRA isoforms, SHANK Associated RH Domain Interactor (SHRPN, SHARPIN), and Epidermal growth factor receptor (ERBB1, EGFR) were significant in both AD datasets, whereas Angiotensin-converting enzyme (ACE), Complement receptor type 2 (CR2), TREM2 and IL34 were significant only in the Bellenguez analysis^9^ (**Figure 5C-D, Supplementary Table 33**). Notably, several of these genes have been implicated in AD GWAS^9^, and the findings are consistent with previous studies highlighting *ACE*, *ERBB1*, *IL34*, *SHRPN*, and *TREM2* as functional candidate genes^24^. Beyond AD, we identified Beta-1,4 N-Acetyl-Galactosaminyltransferase 1 (B4GN1, B4GALNT1) and Ataxin-3 (ATX3, ATXN3) associated with ALS^32,33^, Beta-glucuronidase (BGLR, GUSB)^34,35^ and Syntaxin 6^36,37^ linked to CJD, and ADP-ribosyl cyclase/cyclic ADP-ribose hydrolase 2 (BST1) together with Transmembrane glycoprotein NMB: Extracellular domain (GPNMB) associated with PD^5,38^(**Figure 5E-H, Supplementary Table 33)**.

We explore potential overlapping causal proteins across neurogenerative diseases on the top 100 protein ranking. 10 proteins were shared between AD rankings (Bellenguez-GR@ACE), which is what we were expecting considering that we are investigating the same phenotype. It is remarkable that we observed overlapping between CJD-DLB (10 proteins), AD GR@ACE-ALS (8 proteins), ALS-DLB (8 proteins) and DLB-PD (8 proteins) (**Supplementary Figure 8**). Due to the inclusion of proteins with low-medium reproducibility (12-20%), which could result in biased estimates and results (**Supplementary Figure 9**). The overlaps were also evaluated considering reproducible proteins^30^ revealing similar overlap between DLB-PD, AD rankings, and AD (GR@ACE)-ALS (**Supplementary Figure 10**).

Finally, we investigated whether these MR-informed protein rankings converge on shared biology across neurodegenerative diseases, focusing on reproducible proteins. In DLB ranking, *Vacuolar lumen* (Enrichment Ratio=6.09, FDR=0.004) emerged as prominent signal. Additionally, we identified seven shared mechanisms across diseases, with *Hydrolase activity, acting on glycosyl bonds* representing the most recurrent pathway (PD, DLB and ALS), consistent with a shared pathological axis across these disorders (**Supplementary Figure 11-12**). Importantly, we observed a similar convergence when repeating the analysis considering the complete set of CSF SomaScan measures (**Supplementary Figure 13, Supplementary Table 34-35**).

## Discussion

High-throughput proteomic has transformed our ability to map genetic regulation of protein abundance at scale, enabling systematic discovery of pQTLs across thousands of analytes. In the context of neurodegeneration, profiling the CSF proteome offers a window into pathological processes occurring in the CNS and can illuminate mechanisms underlying diseases such as AD. Here, we characterize the CSF proteogenomic landscape in the ACE CSF cohort, which is deeply phenotyped single-site resource with consensus clinical diagnosis, CSF biomarker-based AT(N) classification, genome-wide genotypes and SomaScan profiles spanning >7,000 aptamer-based measures. To our knowledge, this represents the largest single-site CSF proteogenomic dataset reported to date.

The PCA assessment on SomaScan 7k proteins revealed that several PCs were associated with AD biomarker levels (CSF Aβ42 and p-tau), total protein levels in CSF and the Qalb, which suggests that these factors might be a major contributor to the proteomic variance and should be considered as potential confounders. Previous studies have described multiple significant associations with the AD biomarkers in both CSF and PET imaging, especially with a strongly asymmetric associations with tau pathology^39,40^. Additionally, we observed that the PC1 was explaining the largest proportion of variance in our dataset, which might be driven by the strong correlation among multiple CSF proteins^41–43^.

In this context, we have evaluated several GWAS adjustment models in order to assess the effect of including additional covariates in the number and type of pQTL. The model 3 that included the age, sex, proteomics PC1 and PC2, disease status and 10 genomic PCs as covariates yielded the highest number of observed cis-pQTLs, while minimizing the number of trans-pQTLs. The inclusion of additional covariates might reduce technical bias and promote the detection of robust biological associations. Additionally, we observed that the trans-pQTL located on the chromosome 3 (near *GMNC/OSTN)*, disappeared when adding the proteomic PC1 and PC2 as covariates in the GWAS model 2 and 3. This locus has previously been associated with brain ventricular volume in several studies^44,45^, that might influence CSF proteome levels due to dilution effects caused by increased CSF turnover rates, which could be a confounder for CSF pQTLs^25,43^. Karlsson *et al.*^46^ observed that adjusting for reference CSF proteins, strongly linked to mean standardized protein levels, reduced the number of trans-pQTLs observed in this region. In this context, *GMNC/OSTN* variants have also been associated with tau isoforms (CSF p-tau and t-tau)^44,47,48^. Additionally, we identified 2 well-known trans-pQTLs mapped to the chromosome 6 (*HLA* locus), and 19 (near *APOE*). Consistently, these trans-pQTLs had also been described by Western *et al.* in their analyses^24^, and Hansson et al., identified the *GMNC/OSTN* and *APOE* trans-pQTLs considering an alternative affinity-based proteomic platform, the Olink Proseek® Multiplex panels (CVD-III, INF-I, NEU-I, NEU EXP)^25^.

Notably, the model 3 found the highest number of pQTLs in proteins classified as highly reproducible based on our single correlation score^30,49,50^. These results are also aligned with recent studies comparing affinity and MS proteomics suggesting that pQTL detection by multiple affinity-based techniques provides strong evidence for reduced epitope effects^51^. Moreover, the chi-squared tests confirmed the significant differences in the proportion of pQTLs in reproducible proteins compared to non-reproducible proteins, which supports the validity of our score as key metric for interpreting CSF SomaScan data, emphasizing the importance of conducting a comprehensive quality control of proteome datasets to ensure the reproducibility of proteomic measures strongly influencing downstream results.

Using this cohort, we discovered 264 new loci influencing CSF protein levels and successfully confirmed 511 previously reported genetic markers, with robust concordance across studies. These results support both the robustness of the analytical approach, and the role of these genomic regions as modulators of protein levels. In addition, we identified 80 map refinements (53 cis-, 26 trans-pQTLs) providing finer mapping of previously known loci which could be due to differences in LD structure leading to different lead pQTL findings, and 265 proxy SNPs in strong LD with Western’s pQTLs^24^. Both cis- and trans-pQTLs in these replication categories were mainly linked to mechanisms involved in the immune system and the extracellular matrix organization. Specific cellular conditions might differentially affect gene and protein expression, other studies described specific QTLs active in aging, inflammation, diseases or cell-types^52–54^. Both the innate and adaptative immune system play an important role in pathogen recognition and T-cell regulation. Dysregulation of these mechanisms have been found in several neurodegenerative diseases^55^. Our earlier CSF pQTL atlas highlighted a major HLA trans-pQTL hotspot (chromosome 6) spanning complement-related proteins and implicating immune regulation in CSF protein variation^24^. Concordantly, AD-associated protein candidates prioritised by multiple methods also pointed to immune pathways and cell-types^24^. Other studies have mapped several immune-system and complement cascade genes to the same region^56^. In addition, Sun *et al.* also described a large fraction of trans-pQTLs associated with the complement cascade pathway using plasma samples^57^. Our plasma replications revealed 281 shared CSF pQTLs significant in two independent replication analyses, suggesting that exists systemic modulators of protein levels. These pQTLs were also associated to immune-related and extracellular matrix mechanisms. Moreover, we also found 76 CSF pQTLs exclusively observed in the CNS compartment, which were associated with immune and axon guidance mechanisms.

Conversely, it is well described that co-morbid neuropathological changes can be occurring at the same time than AD, and it might difficult to uncover the extent of the effect of these co-pathologies in cognitive symptoms^58^. Co-pathologies are now being considered in the diagnosis and staging of AD due to its high co-occurrence^59^. To this end, we explored the potential causal effects of the CSF protein levels on AD and other neurodegenerative diseases, using genetic instruments through a MR approach, to identify common molecular pathways or protein modulators. We found both CSF PILRA isoforms (inversely related with AD risk) and CSF SHRPN (directly related with AD risk) as causal proteins for AD in GR@ACE and Bellenguez’s datasets, and CSF ERBB1 was nominally-significant in the GR@ACE analysis, approaching the FDR threshold (inversely related with AD risk). These findings are concordant with the results previously described in literature^24^. Also, *SHRPN* and *ERBB1* genes have been associated to AD in multiple GWAS^8,60^, which is consistent in effect direction with our MR results^24^. Similarly, the downregulation of *ERBB1* gene expression has been related with lower AD risk^9^. In addition, PILRA was nominated as an alternative candidate of AD risk loci in Western’s study, as well as Caro *et al.* finding CSF and plasma PILRA isoforms associated with lower AD risk, reporting a protective association, which is also in agreement with our results^24,61,62^.

Moreover, the AD Bellenguez MR study identified other CSF protein levels that were not found in the AD GR@ACE analysis, including the CR2 (directly related with AD risk), TREM2 (inversely related with AD risk), ACE (inversely related with AD risk) and IL34 (inversely related with AD risk). Previous studies have reported significant elevated CSF TREM2 levels in AD cases^63,64^, with the highest levels observed in MCI due to AD individuals^65^, suggesting that plays a role in AD pathological mechanisms. Also, genetic markers in *TREM2*, and *ACE* have been associated with AD risk in meta-GWAS^7,9,66,67^. Furthermore, *IL34* has been validated in the context of AD in de Rojas *et al.*^8^ and further confirmed in Bellenguez *et al.,*^9^. Additionally, Western *et al*. revealed a significant association between IL34, TREM2 and ACE with AD risk, all these findings are consistent in effect direction as causally linked to AD^24^. In contrast, *CR2* has been proposed as biologically relevant to AD^60^, yet until 2024 it had not been causally implicated via MR using protein-level instruments. Here, we provide convergent evidence supporting a role for CSF CR2 in AD biology. Notably, *CR2* is within the *CR1-CR2* complement receptor cluster on chromosome 1, which harbours one of the most robust AD GWAS loci (*CR1*-rs679515) reported in the largest AD meta-analysis^9^. In our dataset, one *CR2* pQTL showed LD with this *CR1* risk variant (*CR2*-rs6661489 R2=0.884 D’=0.986; *CR2*-rs4844574 R2=0.203 D’=0.503), providing a plausible genetic bridge between the *CR1* locus and CR2 protein regulation. Supporting biological relevance, CSF CR2 levels were inversely associated with p-tau181 (Estimate=-0.06, P=0.015) (**Supplementary Figure 14**). Prior studies are consistent with this axis: Reus *et al.* reported an association between AD risk loci *CR1*-rs3818361 and CSF CR2 levels using Olink^68^, and Western et al. observed colocalized signals linking CSF CR1/CR2 protein levels, and AD risk^24^. Taken together, our results extend this literature by providing protein-instrumented evidence and phenotype anchoring, and suggest that CSF CR2 regulation might contribute to the AD association observed at the *CR1* locus.

Beyond AD, our MR-ready CSF pQTLs inventory revealed several causal associations in other neurodegenerative diseases consistent with previous literature. These include causal associations with concordant effect direction between *BST1*, *GPNMB* and PD (both of which have been described as genetic risk factors)^5,25,38,69–71^. *Syntaxin6* locus was associated with CJD in large GWAS meta-analysis, and multiple studies have supported its association with the disease susceptibility^36,37^. Furthermore, we observed causal associations between *ATX3* (inversely related) and *B4GN1* (directly related) genes in ALS, which previous findings have linked these genes with ALS risk^32,33,72,73^. However, to date no causal association has been found between ATX3 and B4GN1 in ALS, or between Syntaxin6 and CJD. Together, these results support the reliability of our MR findings across multiple neurodegenerative diseases, and highlight potential entry points for therapeutic interventions for protein suppression or supplementation when higher or lower protein levels contribute to disease risk, respectively.

When comparing the overlap of top rankings MR results across neurodegenerative diseases, we observed a modest overlap between PD and DLB rankings in both analyses. This result aligns with the reported genetic correlation among these synucleiopathies^74–76^. When considering only reproducible proteins, we also found an overlap between AD (GR@ACE) and ALS. The genetic correlation between these diseases has also been reported in previous studies and might reflect by common inflammatory and immune-related mechanisms^77^, and comorbid protein deposits.

Furthermore, the *Hydrolase activity, acting on glycosyl bonds* mechanism was observed in the enrichment top ranking of 3 different neurodegenerative diseases, suggesting that metabolic pathways play a general role in the development of neurodegenerative diseases. Importantly, the strong presence of immune-related and inflammatory mechanisms in these enrichment results. The inflammatory processes have been extensively recognised for their implications on pathological mechanisms in neurodegenerative diseases, suggesting a potential detrimental activation of the innate and adaptative immune systems^78,79^. GWAS studies have revealed several loci in immune-related genes associated with an increased AD risk (*CR1*, *HLA*, *TREM2*, *SHRPN*, *IL34* and others^8,9^). These findings support the hypothesis that this process might be a shared pathological mechanism across neurodegenerative diseases^80^. In addition, causal proteins were also enriched in extracellular matrix pathways involving collagen which is the most abundant fibrous protein in the extracellular matrix^81^. Johnson *et al.* observed that CSF proteomic module associated with extracellular matrix was also correlated with AD pathology and cognitive functions^82^, and other components have also been associated with Aβ plaque in AD^83,84^, and α-synuclein aggregates in PD^85,86^.

Despite all these findings, our analyses have several limitations. Although this study represents the largest single site CSF proteogenomic exploration to date, the sample size remains modest compared to large-scale consortia collaborative efforts. The use of an aptamer-based proteomic platform evaluating a fixed number of analytes introduces technical limitations leading to potential platform-specific bias and epitope effects. To address this, we have conducted a comprehensive QC and considered the CSF reproducibility score accounting for several affinity-based methods to ensure the reliability of results^30^. Nevertheless, several SomaScan proteins and genetic markers could not be evaluated for replication in an independent cohort (WashU dataset) due to QC failures. Similarly, MR analysis considering Bellenguez *et al.*^9^ AD meta-GWAS should be considered with caution due to de presence of overlapping samples with our cohort (∼0.26%). To mitigate this issue, we also performed the AD MR analysis considering the fully independent GR@ACE cohort. Finally, limited sample size of several outcome GWAS, such as DLB where no significant results were found, might be limiting the statistical power to detect causal associations compromising the MR analysis^87^. Further studies are needed to elucidate the role of these loci in the modulation of protein levels. Through larger collaborative efforts it would be possible to evaluate the functional relevance and validate these results to disentangle the role of neuroinflammatory processes and extracellular matrix alterations in neurodegenerative diseases.

## Conclusion

In summary, this study represents a large, single-site CSF proteogenomic resource that integrates SomaScan proteomics with genome-wide genotypes to resolve the genetic architecture of the human CSF proteome. We expand the human CSF pQTL landscape by discovering 264 novel loci, independently confirming 511 previously reported associations, and refining known signals through 80 map refinements, and 265 proxy-based replications together providing a high-confidence framework for CSF regulation. By translating these pQTLs into genetic instruments, we further prioritise proteins with evidence consistent with causal roles in AD and other neurodegenerative disorders, thereby nominating mechanistic pathways and candidates for therapeutic development. Beyond the specific loci highlighted here, our results provide a scalable blueprint for linking genetic variation to brain-relevant biology, and a foundation for future functional studies to move from association to mechanisms and, ultimate to intervention.

## Declarations

### Standard protocol approvals, registrations and patient consent

All protocols of the ACE cohort were approved by the Clinical Research Ethics Commission of the Hospital Clinic (Barcelona, Spain, ref. HCB/2014/0494) in accordance with the current Spanish regulations in the field of biomedical research (law 14/2007, royal decree 1716/2011) and the Declaration of Helsinki. In accordance with Spain’s Data Protection Law, all participants

were informed about the study’s goals and procedures by a neurologist before signing an informed consent form. Patient privacy and data confidentiality were protected in accordance with applicable laws. Additionally, the protocols of the HARPONE project were approved by the ethics committee of the Bellvitge University Hospital (Barcelona, Spain, ref. PR067/21).

## Data Availability

The data that support the findings of this study are publicly available from the corresponding authors upon reasonable request. Additionally, the SomaScan proteomic data is publicly accessible through the GNPC and Alzheimer's Disease Data Initiative (ADDI) data-sharing platform (V1 HDS, https://www.neuroproteome.org/).

## Author’s contribution

RP, AC, ACS, and AR designed and conceptualized the study and interpreted the data. RP, AC, ACS and AR contributed to data acquisition, analysis, interpreted the data, and co-wrote the manuscript. PGG, IdR, MCB, CO, AMM, PBB, AV, CY, JT, ML, SC, OSG, BC, AM and FZ contributed to data acquisition and interpretation. AS, LM, MML, MRR, MA, SMG, MVF, MM, SV, PS, XM, LT, MB, SS, MS, CC, AC and AR contributed to data acquisition. RP, AC, ACS, and AR supervised the study. AK, MRR, MA, MVF, MM, SV, JC, PS, XM, LT, BS, MB, SS, MS, CC, AC, ACS and AR contributed to the critical revision of the paper. All authors critically revised the manuscript for important intellectual content and approved the final manuscript.

## Competing interests

All authors declare that the research was conducted in the absence of any conflict of interest.

## Acknowledgements

We would like to thank patients and controls who participated in this project. They were processed following standard operating procedures with the appropriate approval of the Ethical and Scientific Committee. The present work has been performed as part of the doctoral thesis of RPF at the University of Barcelona (Barcelona, Spain).

## Financial support

Authors acknowledge the support of the Agency for Innovation and Entrepreneurship (VLAIO) grant N° PR067/21 for the HARPONE project and the ADAPTED project the EU/EFPIA Innovative Medicines Initiative Joint Undertaking Grant N° 115975. Also, the Spanish Ministry of Science and Innovation, Proyectos de Generación de Conocimiento grants PID2021-122473OA-I00, PID2021-123462OB-I00 and PID2019-106625RB-I00. ISCIII, Acción Estratégica en Salud integrated in the Spanish National R+D+I Plan and financed by ISCIII Subdirección General de Evaluación and the Fondo Europeo de Desarrollo Regional (FEDER “Una manera de hacer Europa") grants PI17/01474, PI19/00335, PI22/01403, PI22/00258 and PI25/00067. The support of CIBERNED (ISCIII) under the grants CB06/05/2004 and CB18/05/00010. The support from PREADAPT project, Joint Program for Neurodegenerative Diseases (JPND) grant N° AC19/00097, and from DESCARTES project, German Research Foundation (DFG). The support of Fundación bancaria “La Caixa”, Fundación ADEY, Fundación Echevarne and Grífols SA (GR@ACE project). AC received support from the Instituto de Salud Carlos III (ISCIII) under the grant Sara Borrell (CD22/00125). PGG is supported by CIBERNED employment plan (CNV-304-PRF-866). IdR is supported by the ISCIII under the grant FI20/00215. JEC received support from National Institute of Health award P30AG066546. Additionally, AR is also supported by STAR Award. University of Texas System. Tx, United States, The South Texas ADRC. National Institute of Aging. National Institutes of Heath. USA. (P30AG066546), the Keith M. Orme and Pat Vigeon Orme Endowed Chair in Alzheimer’s and Neurodegenerative Diseases and Patricia Ruth Frederick Distinguished Chair for Precision Therapeutics in Alzheimer’s and Neurodegenerative Diseases.

## Abbreviations

AD: Alzheimer’s disease; ALS: Amyotrophic lateral sclerosis; ANML: Adaptive normalization by maximum likelihood; APOE : Apolipoprotein E; AT(N): Amyloid, Tau, Neurodegeneration classification; Aβ: Amyloid β; Aβ42: Amyloid beta 42 peptides; BBB: Blood Brain Barrier; CDR: Clinical Dementia Rating; CJD: Creutzfeldt-Jakob disease; CNS: Central nervous system; CSF: Cerebrospinal fluid; CV: Coefficient of variation; D’: Normalized coefficient of linkage disequilibrium; DLB: Dementia with Lewy-bodies; DLB: Dementia with Lewy-bodies; DNA: Deoxyribonucleic acid; ELISA: Enzyme Linked Immunosorbent Assay; ECM: extracellular matrix; FDR : False discovery rate; FUMA: Functional Mapping and Annotation of GWAS; GWAS: Genome-Wide Association Studies; HC: Healthy controls; ID: Identification; INDELs: insertions and deletions; IQR: Interquartile range; IVW: Inverse variance weighted P-value; kb: Kilobase pairs; KEGG: Kyoto Encyclopedia of Genes and Genomes; LD: Linkage disequilibrium; LP: Lumbar puncture; MAC: Minor allele count; MAF: Minor allele frequency; Mb: Megabase pairs; MCI: Mild Cognitive Impairment; Meff: Effective number of independent tests; MMSE : Mini-Mental State Examination; MR: Mendelian Randomization; MR-PRESSO: Mendelian Randomization Pleiotropy RESidual Sum and Outlier; MS: Mass spectrometry; NBACE: Neuropsychological battery of ACE; ORA: Over-Representation Analysis; PC: Principal component; PC1: First principal component; PC2: Second principal component; PCA : Principal components analysis; PD: Parkinson’s disease; pQTL: Protein quantitative trait loci; p-tau: Hyperphosphorylated Tau at Thr 181; Q1: First quantile; Q3: Third quantile; Qalb : Albumin quotient index; QC : Quality Control; R2: Imputaion quality; R²: Allelic correlation coefficient; RFU: Relative fluorescent units; SCD: Subjective cognitive decline; SD: standard deviation; SNAP: Suspected Non-Amyloid Pathology; SNP: Single nucleotide polymorphism; TMT: Tandem mass tag; TSS: Transcription start or end position site; t-tau: Total Tau; UniProtID: UniProt Swiss-Prot identifier codes.

## Materials and Methods

### Study participants and Sample Collection

ACE is a private non-profit foundation with a specialized memory clinic with extensive experience in the care of patients with cognitive decline. Over time, at ACE we have evaluated more than 34,300 patients, collected and analysed 25,000 genetic samples, and participated in over 165 clinical trials. The clinical diagnosis of each patient is established by a multidisciplinary group of clinicians, neuropsychologists and social workers. The ACE cohort consists of individuals across the entire cognitive spectrum, classified into the following categories: 1) HC and 2) subjective cognitive decline (SCD)^88^ both with a Clinical Dementia Rating (CDR)^89^ of 0; MCI diagnoses that were assigned to a CDR of 0.5, the neuropsychological battery of ACE (NBACE)^90^ accounting for the cut-offs of impairment for age, formal education levels, the classification of López *et al.* 2003 and Petersen’s criteria were also considered^88,91–94^. Additionally, the 2011 NIA-AA guidelines were used to establish the AD dementia diagnosis^29^. Clinical characterization of these individuals has been described elsewhere^95,96^.

A lumbar puncture (LP) was performed to obtain CSF and measure CSF AD-related biomarkers, following the consensus recommendations^97^. We collected the CSF samples, centrifuged (2000xg 10 min at 4°C) and stored at -80°C. For CSF Aβ42 and p-tau protein determination, an aliquot was defrosted at room temperature and vortexed. Protein levels were measured using the commercial Enzyme-Linked ImmunoSorbent Assay (ELISA) kits Innotest β-AMYLOID (1-42) and chemiluminescent enzyme-immunoassay Lumipulse G600II automated platform (Fujirebio Europe, Göteborg, Sweden)^95^. Peripheral blood was collected to obtain DNA using the Chemagic System (Perkin Elmer) and Maxwell RSC48 (Promega) following standard procedures. DNA samples with a high integrity and concentrations higher than 10 ng/μL were selected for genotyping.

### SOMAScan proteomic QC and Processing

We analysed 1,370 paired plasma and CSF samples, corresponding to 1,325 individuals, using the aptamer-based proteomic platform SomaScan 7k Assay version 4.1 (SomaLogic Operating Co., Inc. Boulder, Colorado). This platform measures 7,596 unique aptamers corresponding to 6,402 distinct human proteins based on unique UniProt identification codes (UniProtID). Briefly, this assay uses a 50 µL CSF sample and modified DNA aptamers to measure protein abundance detected using fluorescence in a conventional DNA array as described elsewhere^14^. Finally, the protein abundance was expressed in relative fluorescent units (RFU) and normalized using the adaptive normalization by maximum likelihood (ANML) method^98,99^. We used multiple CSF aliquots in the first thawing cycle for conducting these proteomic analyses.

Quality control (QC) was performed using the WashU in-house protocol^24^. In brief, we removed aptamers following either of these criteria: 1) the maximum absolute difference between the aptamer scale factor and the median scale factor if any plate higher than 0.5, and 2) the median coefficient of variation (CV) between plates higher than 15%. The remaining proteomic measures were log10-transformed and proteomic values outside the 1.5-fold of the interquartile range (IQR) were considered outliers and excluded from further analysis, values higher than the 1.5-fold IQR + third quantile (Q3) and values lower than 1.5-fold IQR - first quantile (Q1). Then, we removed aptamers and samples with a call rate lower than 65%. Subsequently, we recalculated the call rate and removed first aptamers and then samples at a more restrictive threshold (call rate<85%). We then standardized protein values using the R function *scale* with both *center* and *scale* set to TRUE. Prior to the QC, ACE cohort included 7,596 aptamers measured in 1,370 samples. For the aptamer QC steps, there were 72 aptamers that failed the median scale factor (Criteria 1), and 148 aptamers removed in the inter-plate coefficient of variation (Criteria 2) step. Of these, 56 had failed both criteria. At this point there were 7,376 aptamers measured in 1,370 individuals remaining for further analysis. After the log10-transformation, we removed 253,063 outlier proteomic measures through the IQR. In evaluating the call rate, there was 1 aptamer that failed the call rate criteria 85%. Furthermore, we removed 11 and 29 individuals in the sample call rate filtering at 65% and 85%, respectively. We also removed non-human and non-protein aptamers (n=283), longitudinal samples by selecting one for each individual, and samples with no genomic information (n=53). After these proteomic QC pipeline, there were 8,901,309 proteomic measures from 7,092 SomaScan aptamers measured on 1,277 individuals available for further analyses (**Supplementary Figure 15, Supplementary Table 36-37**).

### Principal components analysis (PCA)

Considering the proteomic data that passed the QC, we performed a principal component analysis (PCA). Again, the information of each individual was also scaled as described above. We computed the covariance matrix using the *cov* R function using *pairwise complete observations*. We then conducted the PCA using the *princomp* function for obtaining the principal component (PC) information using the R version 4.3.1. In addition, we evaluated the number of PCs required to explain the 95% of variance (n=8), and we analysed the contribution of multiple clinical phenotypes to the first 5 PCs. These included age, sex, CSF AD biomarkers, BMI, CSF total protein levels, Qalb, *APOE* genotype, disease status and AT(N) classification [Amyloidosis, Normal, Prodromal AD, and Suspected Non-Amyloid Pathology (SNAP)], using linear associations and non-adjusted Pearson correlations.

### SomaScan reproducibility score in CSF

In our previous work, we created a reproducibility metric for CSF SomaScan proteomic data based on Spearman correlation comparing two independent SomaScan assays, and comparing with the antibody-based proteomic platform Olink Explore, as described elsewhere^30^. We used multiple aliquots of the same samples to conduct these proteomic analyses, and integrated these correlation results into a 12-category reproducibility score (Single.Score.Correlation). We considered the aptamers included in the score categories 1-3 and A as reproducible SomaScan measures in CSF^30^ (**Supplementary Table 2**).

### Genomic QC and Data processing

DNA samples from the Ace cohort were genotyped using the Axiom 815K Spanish Biobank Array (Thermo Fisher) at the Spanish National Center for Genotyping (CeGEN, Santiago de Compostela, Spain). In brief, individuals with low-quality samples, excess of heterozygosity, sample call rate below 97%, sex discrepancies, variants call rate below 95% or a deviation from the Hardy–Weinberg equilibrium (P>1e-06) were excluded from the analysis, as described elsewhere^8,96^. The imputation was performed using the TopMed imputation server^100^ (Michigan, USA). Only variants with a MAF>1%, a minor allele count (MAC)=10 and high imputation quality (R2≥0.3) were kept for the subsequent analyses. To account for population structure and detect non-European population outliers, we calculated the PCA using PLINK 1.9 and removed individuals deviating from the mean PC of the European population (>3SD). Also, we removed related individuals though an IBD analysis (PI_HAT≥0.1875). Briefly, we excluded 12 individuals due to cryptic relatedness, 5 individuals due to non-European ancestry and 1 individual due to both reasons (**Supplementary Figure 16**). Finally, there were 1,259 individuals with available genomic information that passed the quality control were considered for analysis, including 9,016,686 SNPs after imputation (**Supplementary Table 1**).

### pQTL identification

For pQTL identification, we first conducted a PCA using pre-imputed genotyping data to account for population microstratification in the GWAS. We then conducted an additive generalized linear model using the *--glm* flag in PLINK 2 software and analysed 3 models considering the following covariates: A) considering the age, sex and 10 genomic PCs (Model 1, Equation 1)^24^, B) considering the first and second proteomic PCs (pPC1 and pPC2) (Model 2, Equation 2), and C) considering the age, sex, pPC1 and pPC2, disease status as dummy variables and 10 genomic PCs (gPC) (Model 3, Equation 3). The models follow the equations below, where Y denotes the outcome variable (aptamer measures), β refers to regression effect sizes including both β_i_ and β_j_, which refer to the regression effect sizes of gPCs and pPCs respectively, and ε to the error term.

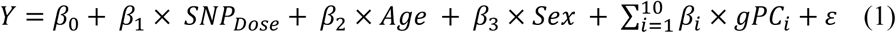

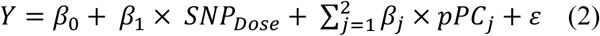

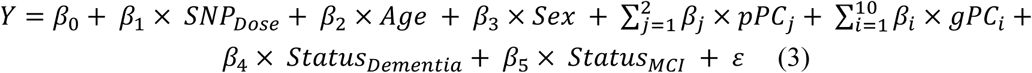

The genomic inflation factor or lambda (λ) was computed for each GWAS models, and suggested no gross bias or stratification in these analyses (**Supplementary Figure 17**).

#### Cis- and trans-pQTL classification

For classifying the pQTLs into cis- or trans-pQTLs, we harmonized the information obtained from the GENCODE v46 and UniProt tables as described below. We then converted the chromosomal positions to kilobase pairs (kb) and computed the absolute positions. We defined as cis-pQTLs as those significant genetic markers (P<5e-08) associated with protein levels that were mapped at a 1 megabase paire (Mb) window of the transcription start site (TSS) of the gene encoding for that protein (±500 kilobase pairs (kb) upstream/downstream the TSS). Conversely, we defined the trans-pQTLs with a more stringent significance threshold, considering genome-wide significant genetic markers at P<6.25e-09 associated with protein levels outside 1 Mb window from the TSS (P<5e-08 / n, n=8, PCs needed to explain 95% of variance)^24^. We also indicate if protein could not be mapped to any genomic regions mainly because some aptamers are targeting protein complex that have more than one UniProtID.

### Gene-Protein annotations

The genomic regions were downloaded from the TableBrowser tool^101^ to export data from the University of California Santa Cruz (UCSC) Genome Browser annotation track database (https://genome.ucsc.edu/cgi-bin/hgTables). To download the data, we filtered the human genome in the GRCh38/hg30 (Dec. 2013) assembly and selected the Genes and Gene prediction group. We selected the table *knownGene* from the track *GENCODE V46*, to download the complete genome region. Also, we included in the table the complete set of *hg38.knownGene* and *hg38.kgXref* variables. We also downloaded the protein information from TableBrowser by selecting the *SwissProt Aln. (unipAliSwissprot)* table from the *UniProt* track. We included the chromosomal coordinates for these proteins in the GRCh38/hg30 (Dec. 2013). We integrated the information included in the GENCODE and UniProt datasets selecting the protein primary sequence to provide chromosomal positions for each of the UniProtID.

### pQTL replications

In addition, we replicated CSF pQTL findings in a fully independent pQTL analysis conducted using the WashU dataset^24,102^, considering the model 3 covariates and the genotyping array as dummy variable. The CSF pQTL replication analysis included 1,759 SomaScan aptamers passed QC (7k platform) and 2,784 individuals from the cohorts ADNI, DIAN, Knight-ADRC (October 2021) and Knight-ADRC (June 2023), Barcelona-1, PPMI, Stanford ADRC and Stanford Aging Memory Study, MARS. The samples from the ACE CSF cohort were excluded. The significance threshold for replication was set to P<0.05. Additionally, to evaluate if pQTL findings had been described in previous literature, we considered the pQTL list reported by the largest pQTLs analyses conducted in CSF samples to date by Western *et al*. (n=2,477)^24^.

Based on the CSF pQTL replication data, we classified the pQTL into 4 categories: 1) Novel CSF pQTLs, including significant variants in the independent replication (WashU) that were not reported by Western *et al.*^24^, 2) Replicated CSF pQTLs, which were found significant across the three studies: ACE CSF analysis, replicated in an independent pQTL analysis (WashU), and had been previously reported^24^, 3) Proxy SNPs in high LD (R^2^≥0.5) with a pQTL described in Western et al. at 1Mb window; and 4) Map refinements where we found a more informative marker at a previously known locus. For replicated pQTLs, we compared the effect size (betas coefficients) from our study with those reported by WashU independent replication, and we evaluated the LD (R^2^ and D’) between novel pQTLs, and pQTLs reported in Western’s analyses^24^ within a ±1Mb window around each lead variant. LD estimation was conducted using PLINK2 on non-overlapping control samples from the GR@ACE cohort (n=8,475). To further characterize this subset of novel CSF pQTLs, we conducted a conditional GWAS considering model 3 covariates and the pQTL described by Western et al^24^. Additionally, we investigated the biological mechanisms associated with the pQTLs classification using the Over-Representation Analysis (ORA) WebGestalt online tool (https://www.webgestalt.org/)^103^. We have considered the *Homo sapiens* as Organism, *Gene/Protein* as Analyte Type, the *dbSNP* identification codes and we used the *genome* as reference list. We have considered multiple functional databases for the Over-Representation Analysis (ORA) (**Supplementary Table 38**).

Furthermore, we conducted a plasma pQTL replication to determine whether CSF pQTLs were systemic modulators of protein levels, also influencing protein abundance in plasma, or were exclusive of the CNS. For the plasma replication we used 2 datasets: 1) replication A, considering the plasma WashU dataset including 1,657 SomaScan aptamers passed QC (7k platform) and 2,338 individuals from the Knight-ADRC cohort. Further details can be found elsewhere^24,102^; and 2) replication B, corresponding the plasma pQTL evaluation conducted by Pietzner *et al*^20^.

For replication A, we assessed concordance between CSF sentinel (genome-wide significant, LD independent) cis-pQTLs and plasma cis-pQTLs considering the WashU dataset. We excluded insertions and deletions (INDELs), harmonized effect alleles and effect size. We also evaluated significance in plasma considering a P-value threshold correcting by the 755 unique proteins. For replication B, 1,281 CSF cis-pQTLs (excluding INDELs) were considered, plasma pQTL GWAS summary statistics were obtained from Pietzner *et al*.^20^, which profiled 4,775 proteins in 10,708 European-descent participants from the Fenland study using the SomaScan v4 assay^20^. Genomic coordinates were lifted over to GRCh37 using *pyliftover* to match the plasma dataset. For each CSF sentinel variant and the corresponding protein, we extracted plasma GWAS results for variants within ±250kb window. Within each locus, we controlled for multiple testing using the Genetic Type I error calculator (GEC)^104^ to estimate the effective number of independent tests (Meff), accounting for the linkage disequilibrium (1000 Genomes Project Phase 1 version 3, European ancestry, GRCh37 build) and set a locus-specific threshold (P_GEC_=0.05/Meff). A locus was considered concordant if any plasma variant within ±250 kb less than P_GEC_. We then aligned effect alleles and harmonized effect sizes. After harmonization, 645 CSF cis-pQTLs had allele-aligned plasma counterparts that met a significance criterion (both P_CSF_, P_plasma_<P_GEC_/532), correcting for the 532 unique proteins tested. Finally, a Kyoto Encyclopedia of Genes and Genomes (KEGG) pathway enrichment analysis was performed for both replication analyses on the overlapping proteins between CSF and plasma cis-pQTLs using the GENE2FUNC module of the Functional Mapping and Annotation (FUMA) of GWAS platform^105^.

### Mendelian randomization analysis

We selected independent genome-wide significant cis-pQTLs as an instrument variant for the MR analyses. We perform the clumping of genetic variants with P<5e-08 for the index SNP (*--clump-p1*), secondary significance threshold of 1e-5 (*--clump-p2*), a linkage disequilibrium (LD)-threshold of 0.001 (*--clump-r2*) at a physical distance of 250 kb (*--clump-kb*) to select independent genomic variants using the PLINK1.9 software. We excluded from further analysis generic markers located in pleiotropic regions defined by Western el al^24^, 2 cis-pQTLs on the chromosome 3 (*GMNC*/*OSTN)*, 39 cis-pQTLS on the chromosome 6 (*HLA* locus), and 1 cis-pQTL on the chromosome 19 near *APOE.* We also evaluated the pQTL strength for MR analyses by calculating the F-statistic using the formula (4)^106^, all SNPs had sufficient strength (F>10, range: 30.12-9439.32) and were considered genetic instruments.

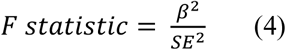

Additionally, we downloaded the GWAS summary statistics of neurodegenerative diseases to be considered as an outcome in the MR analyses. including AD, PD^5^, CJD^36^, DLB^4^, and ALS^4^. For the AD, we conducted an independent GWAS using the GR@ACE cohort considering 7,062 cases and 8,474 controls of European ancestry following our established in-house procedure described elsewhere using the PLINK2 software^8^. Alternatively, we also analysed the summary statistic of the largest AD GWAS to date (Bellenguez’s study)^9^.

We harmonized the GWAS summary statistic and then evaluated whether the selected cis-pQTLs were represented in the outcome summary statistics. If the genetic marker was not represented in the outcome GWAS, we searched proxy SNPs based on correlation coefficient (R^2^), normalized coefficient of linkage disequilibrium (D’) and genomic distance using the *LDproxy* function from the *LDlinkR* package considering the CEU population of European ancestry. Genetic markers with R^2^<0.5 were considered as different SNPs.

We conducted the MR analysis for each CSF SomaScan aptamer using 6 methodologies to evaluate the potential causal effect of protein levels on the aetiology of neurodegenerative diseases. We used the *mr_allmethods* function of the *MendelianRandomization* R package, specifying the method *main*. For the aptamers that only had one instrument marker, the *inverse variance weighted P-value* (IVW) method simply considers the Wald ratio method for analysis. Conversely, for proteins that had more than 2 instrument markers, several MR methodologies were used including simple and weighted median, IVW and Egger methods. We performed additional MR strategies on proteins that had more than 2 genetic instruments: 1) Contamination mixture method was performed using the *mr_conmix* function of the *MendelianRandomization* package, CIMin=-1 and CIMax=1, and 2) Leave One Out method was conducted using the *mr_loo* function of the *MendelianRandomization* package. Lastly, the Mendelian Randomization Pleiotropy RESidual Sum and Outlier (MR-PRESSO)^107^ was conducted using the *mr_presso* function of the *MRPRESSO* package, considering the OUTLIERtest and DISTORTIONtest set to TRUE, NbDistribution=1000.

Lastly, considering the top 100 ranked protein from the MR results of neurodegenerative diseases, we investigated in which molecular mechanisms were involved these proteins using the WebGestalt tool^103^. We have followed a similar pipeline as described above, using UniProtID and the custom reference list accounting for the 7,092 SomaScan proteins that passed the proteomic QC.

## Notes

### Competing Interest Statement

The authors have declared no competing interest.

### Author Declarations

All protocols of the ACE cohort were approved by the Clinical Research Ethics Commission of the Hospital Clinic (Barcelona, Spain, ref. HCB/2014/0494) in accordance with the current Spanish regulations in the field of biomedical research (law 14/2007, royal decree 1716/2011) and the Declaration of Helsinki. In accordance with Spain's Data Protection Law, all participants were informed about the study's goals and procedures by a neurologist before signing an informed consent form. Patient privacy and data confidentiality were protected in accordance with applicable laws. Additionally, the protocols of the HARPONE project were approved by the ethics committee of the Bellvitge University Hospital (Barcelona, Spain, ref. PR067/21).

