## Supplementary Figures for "Human CSF proteogenomics links genetic variation to neurodegenerative disease proteins"

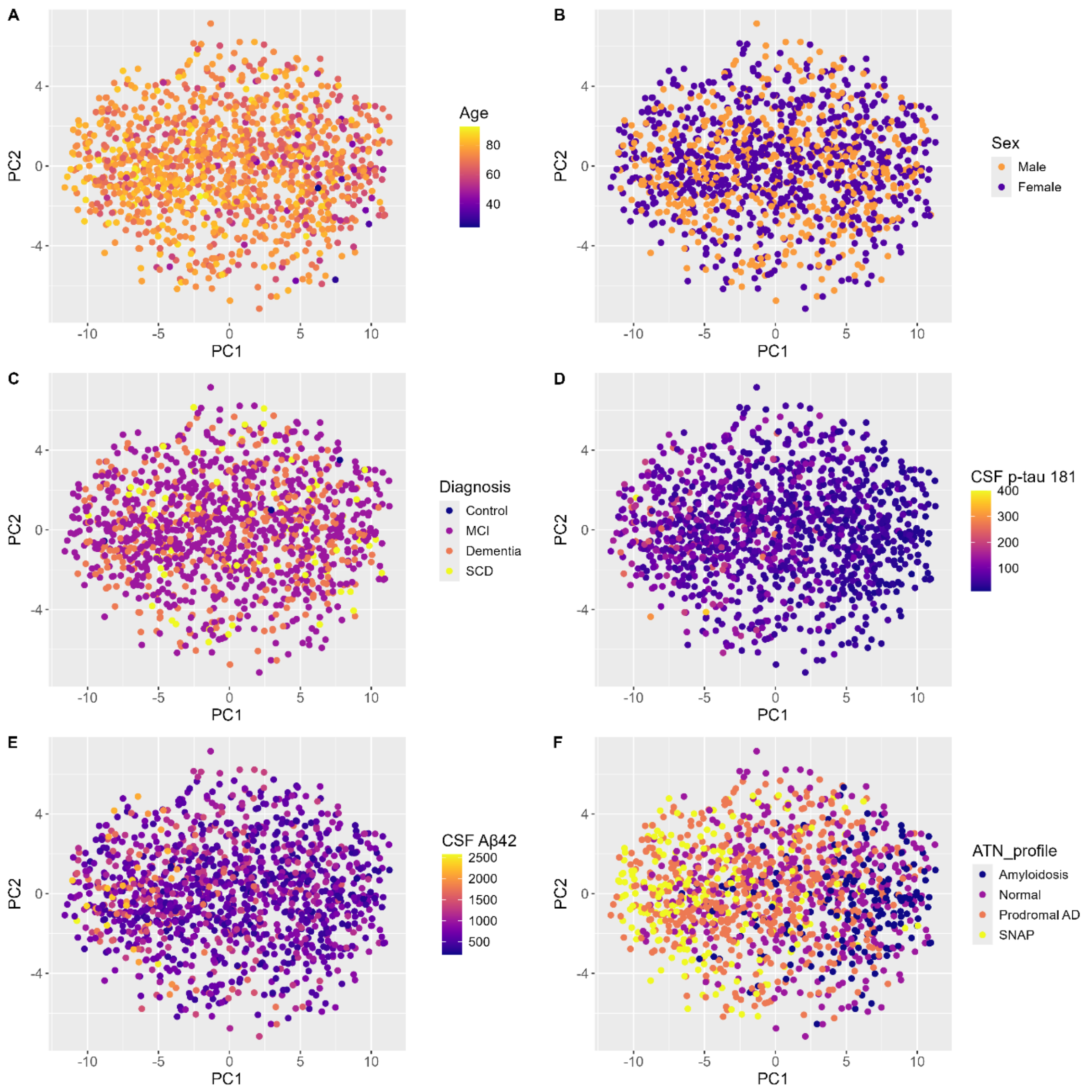


***Supplementary Figure 1. Graphical representation of PC1 and PC2 variables coloured by multiple CSF biomarkers, clinical and demographic variables.*** *A) Age, B) Sex, C) Disease Status, D) CSF p-tau 181, E) CSF Aβ42 and F) ATN profile.*

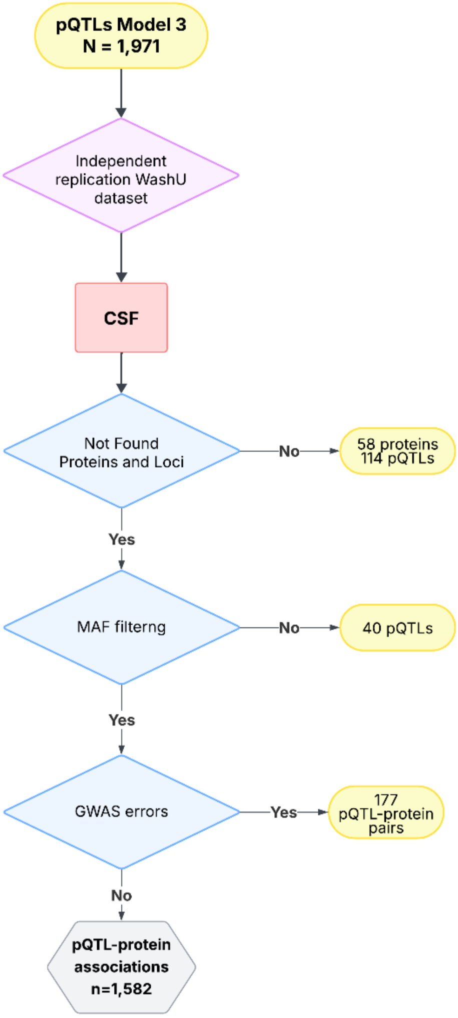


***Supplementary Figure 2. Flow diagram of the independent replication of ACE CSF pQTLs (model 3) considering the WashU dataset.*** *Description of the CSF proteins-loci included in the WashU independent replication where the ACE samples were excluded.*

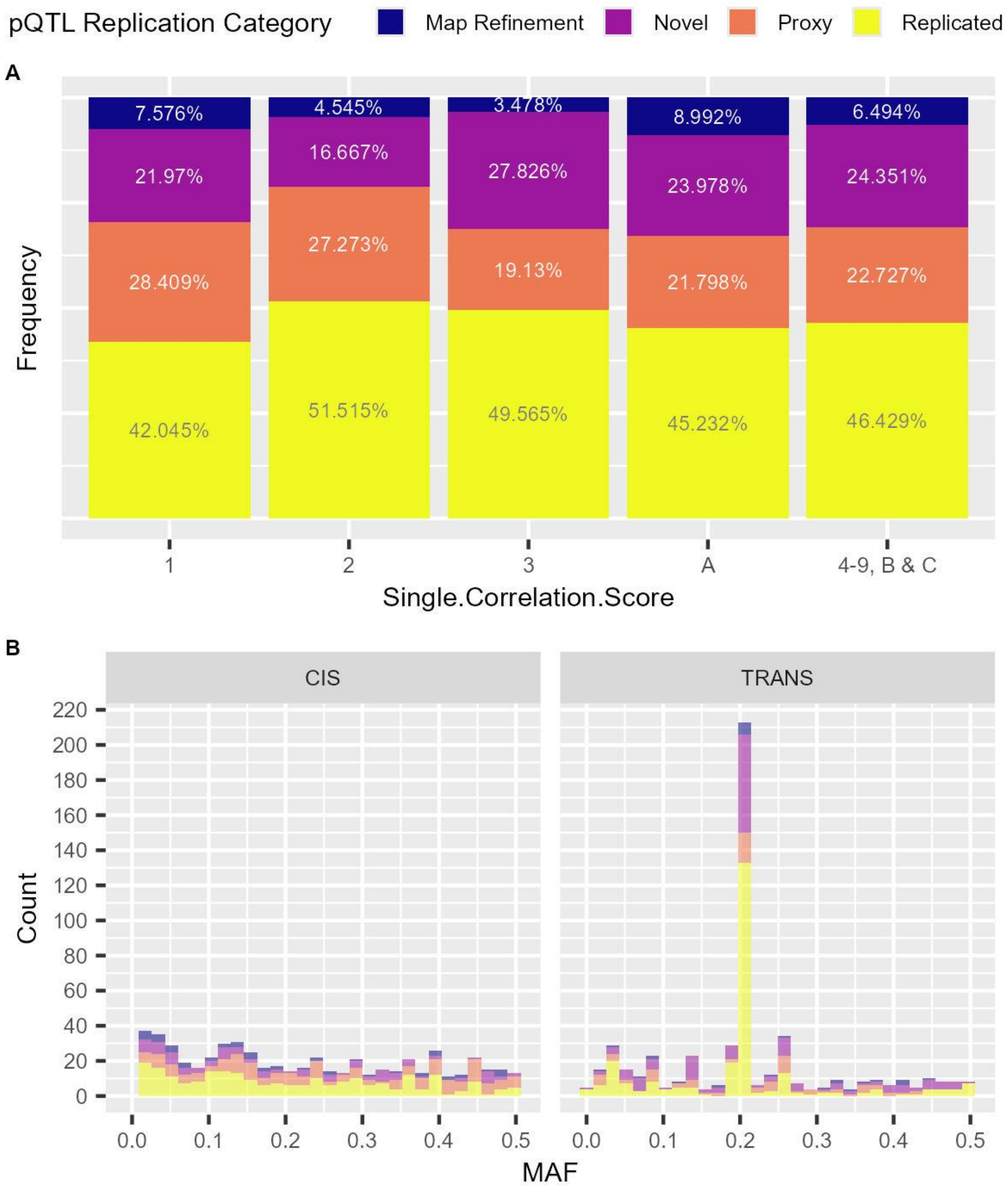


***Supplementary Figure 3. Exploration of the CSF pQTL replication results.*** *A) Distribution of the reproducibility score in the model 3 pQTLs coloured by the replication categories of the comparison with Western et al., 2024. B) Stacked bar plot of the MAF distribution across cis- and trans-pQTLs coloured by replication category.*

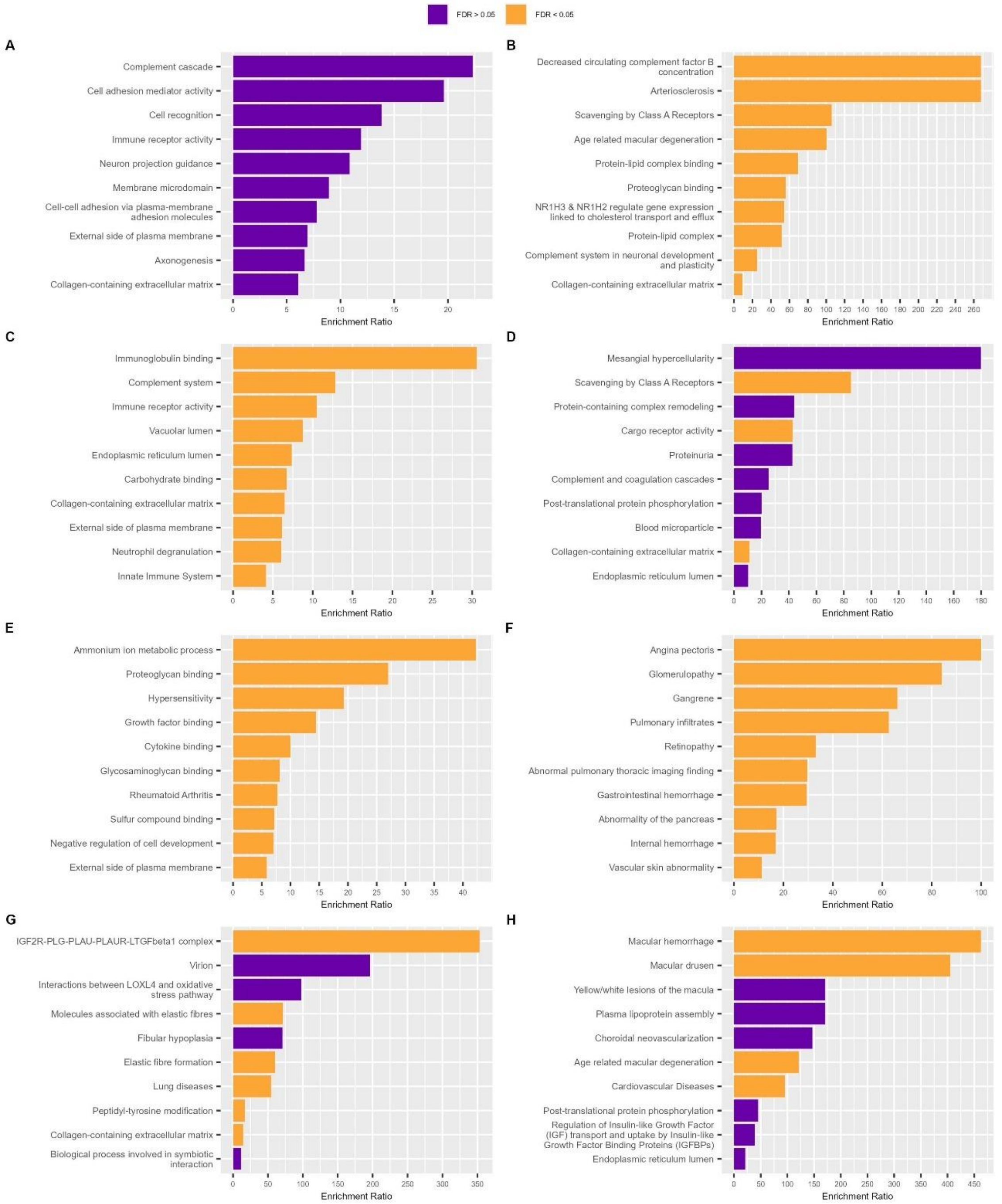


***Supplementary Figure 4. Top 10 enriched mechanisms of the replication categories in CSF including only reproducible SomaScan proteins.*** *We conducted an ORA analysis using the WebGestalt tool (WG). A) Enriched mechanisms of novel cis-pQTLs, B) Enriched mechanisms of novel trans-pQTLs, C) Enriched mechanisms of replicated cis-pQTLs. D) Enriched mechanisms of replicated trans-pQTLs. E) Enriched mechanisms of proxy cis-pQTLs. F) Enriched mechanisms of proxy trans-pQTLs. G) Enriched mechanisms of map refinement cis-pQTLs, H) Enriched mechanisms of map refinement trans-pQTLs.*

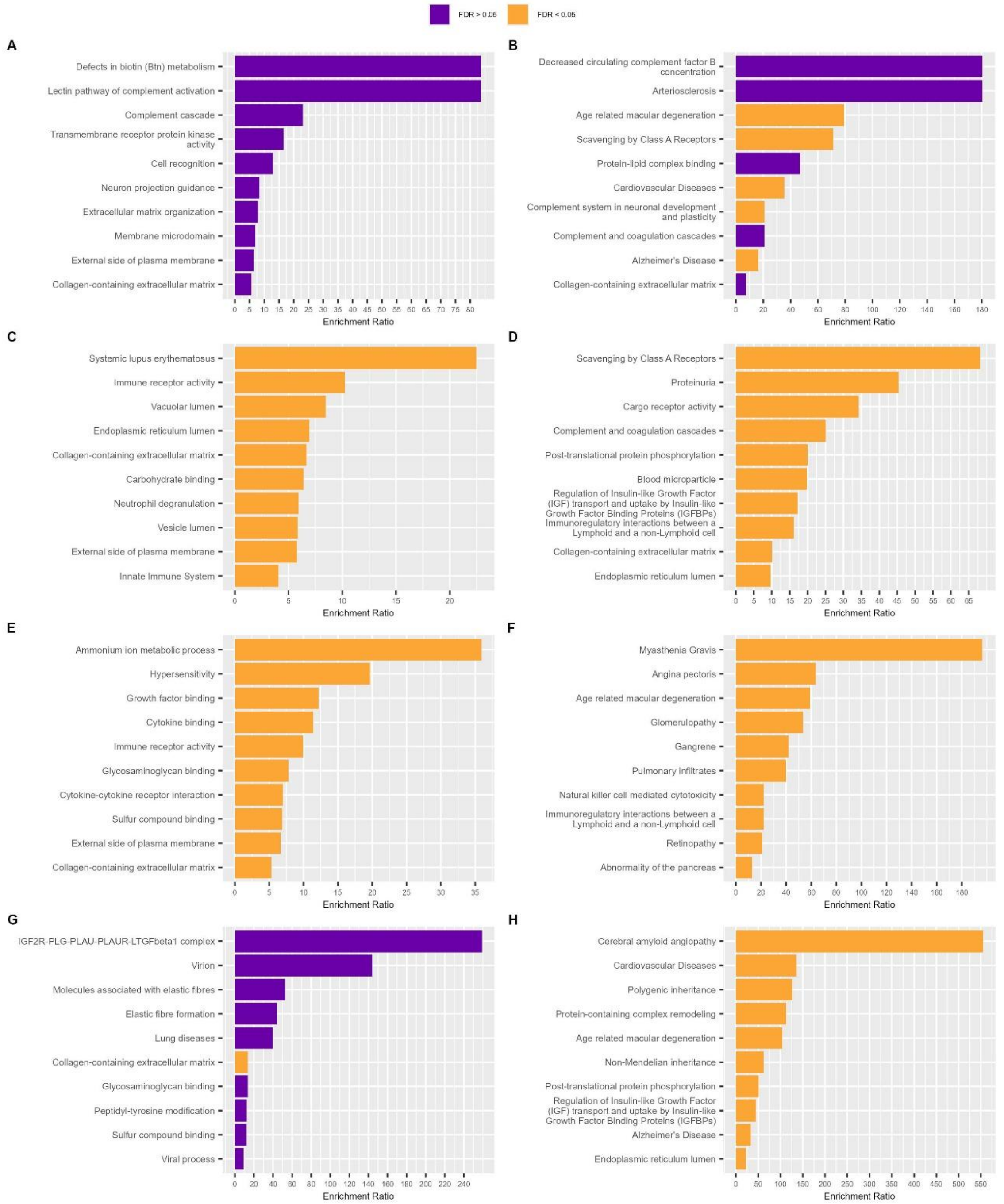


***Supplementary Figure 5. Top 10 enriched mechanisms of the replication categories in CSF considering the complete set of SomaScan proteins.*** *We conducted an ORA analysis using the WebGestalt tool (WG). A) Enriched mechanisms of novel cis-pQTLs, B) Enriched mechanisms of novel trans-pQTLs, C) Enriched mechanisms of replicated cis-pQTLs. D) Enriched mechanisms of replicated trans-pQTLs. E) Enriched mechanisms of proxy cis-pQTLs. F) Enriched mechanisms of proxy trans-pQTLs. G) Enriched mechanisms of map refinement cis-pQTLs, H) Enriched mechanisms of map refinement trans-pQTLs.*

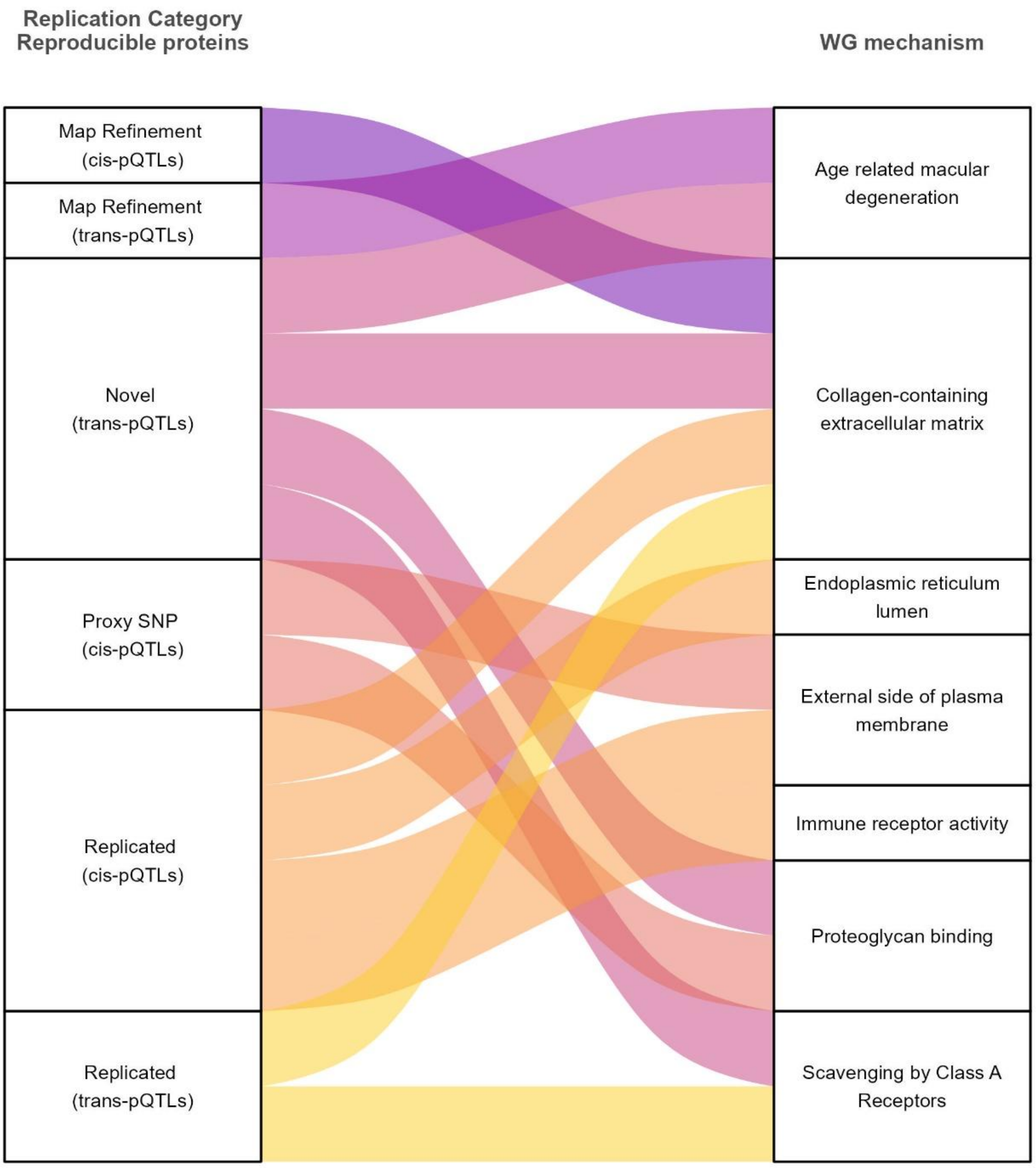


***Supplementary Figure 6. Enriched mechanisms identified across all replication categories between ACE and Western et al., 2024 analyses.*** *Representing FDR-significant mechanisms considering cis- and trans- pQTLs in reproducible proteins. The WebGestalt tool (WG) was used for these analyses.*

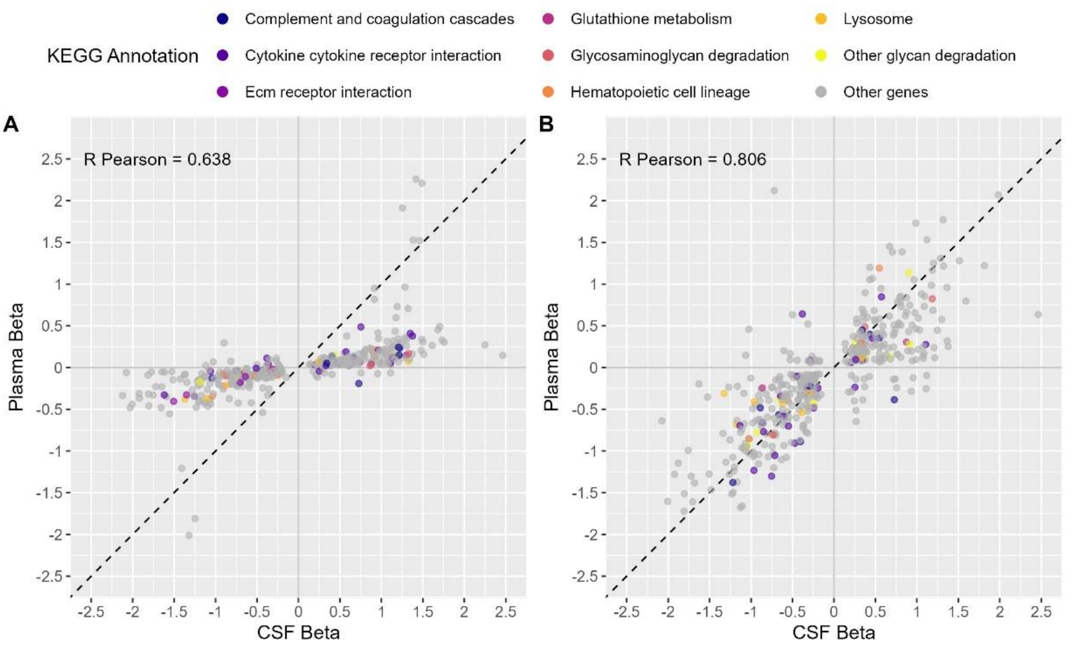


***Supplementary Figure 7. Exploration of the cis-pQTLs independent replication in plasma.*** *A) Scatterplot of beta (effect size estimate) between the ACE CSF model 3 and the plasma cis-pQTL exploration from the replication A (n=438). B) Scatterplot of beta (effect size estimate) between the ACE CSF model 3 and the plasma cis-pQTL exploration from the replication B (n=421). Both comparisons were coloured considering the KEGG annotations obtained by FUMA GENE2FUNC tool ([https://fuma.ctglab.nl/](https://fuma.ctglab.nl/)).*

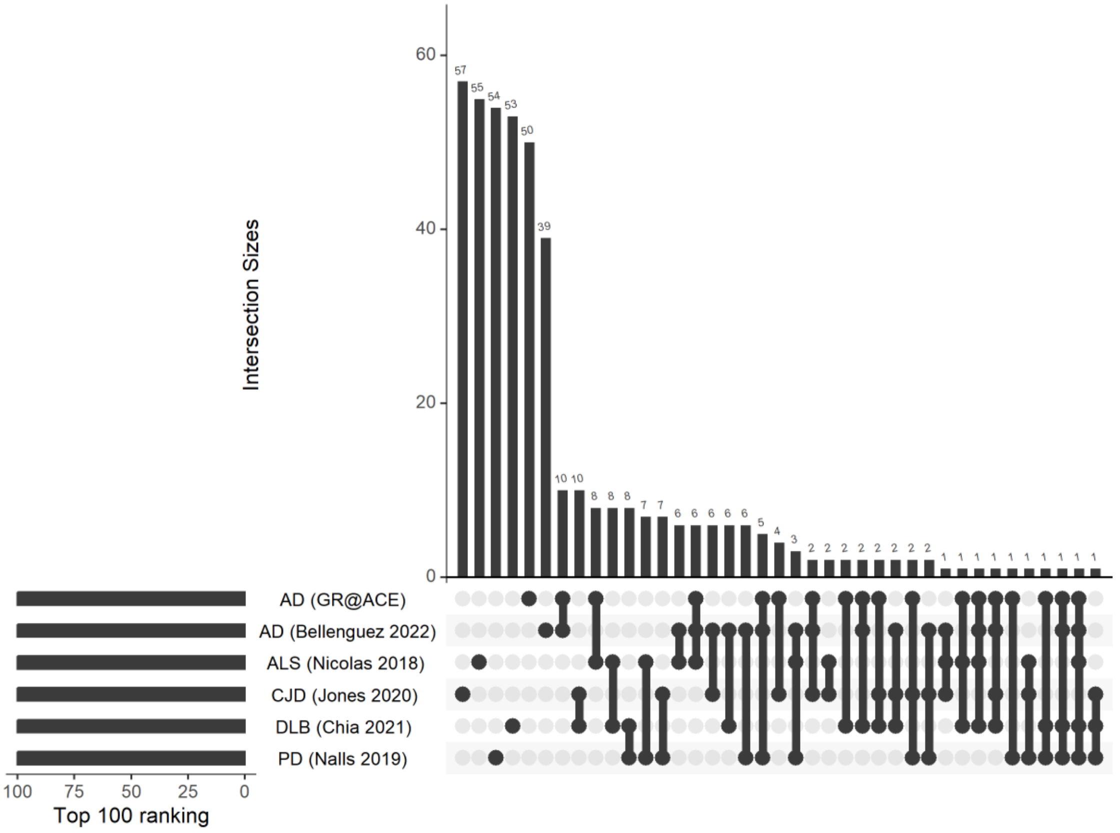


***Supplementary Figure 8. Overlapping results between FDR significant proteins included in the top 100 ranking of neurodegenerative MR results****. All proteins were included.*

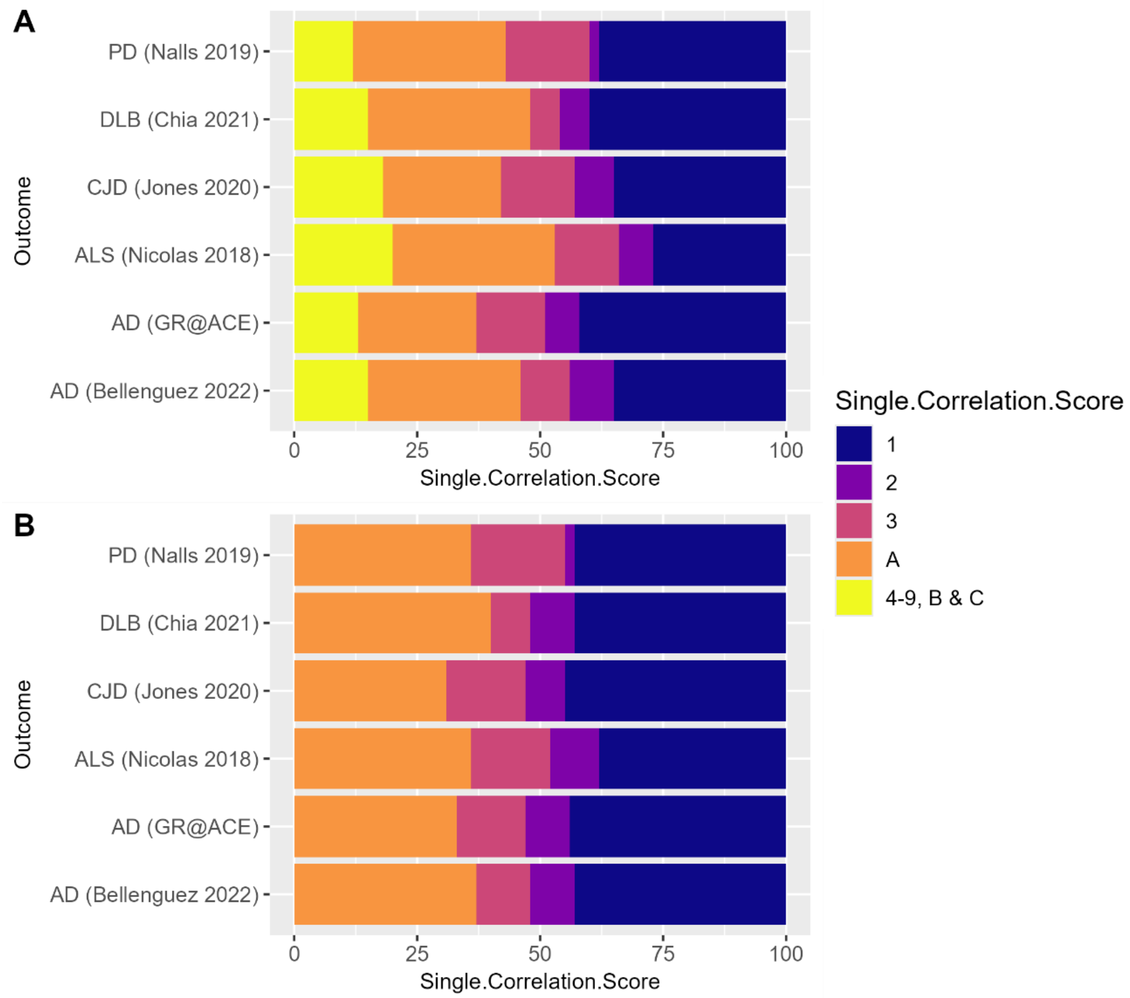


***Supplementary Figure 9. Distribution of the Reproducibility score in each neurodegenerative disease top 100 ranking.*** *A) Reproducibility Score considering the complete set of proteins. B) Reproducibility Score considering only reproducible and reliable proteins.*

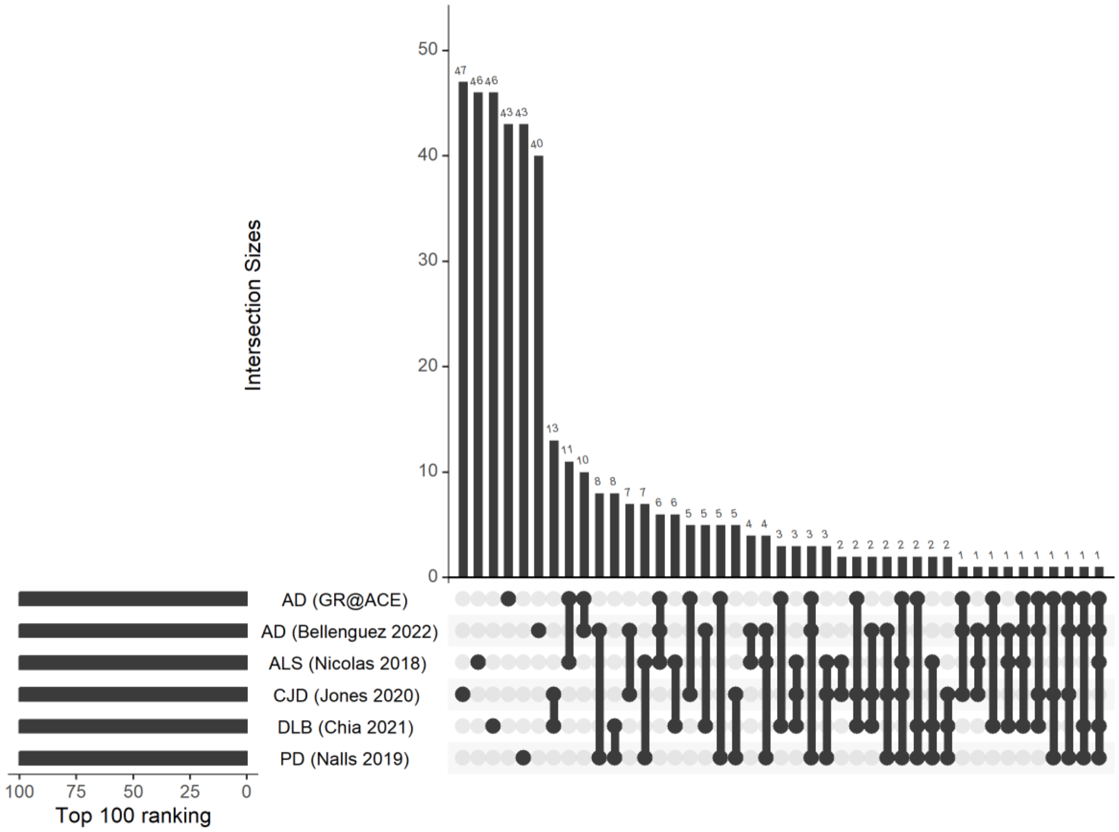


***Supplementary Figure 10. Overlapping results between FDR significant proteins included in the top 100 ranking of neurodegenerative MR results.*** *Only reproducible proteins, represented in the score categories 1-3 and A, were included for these analyses.*

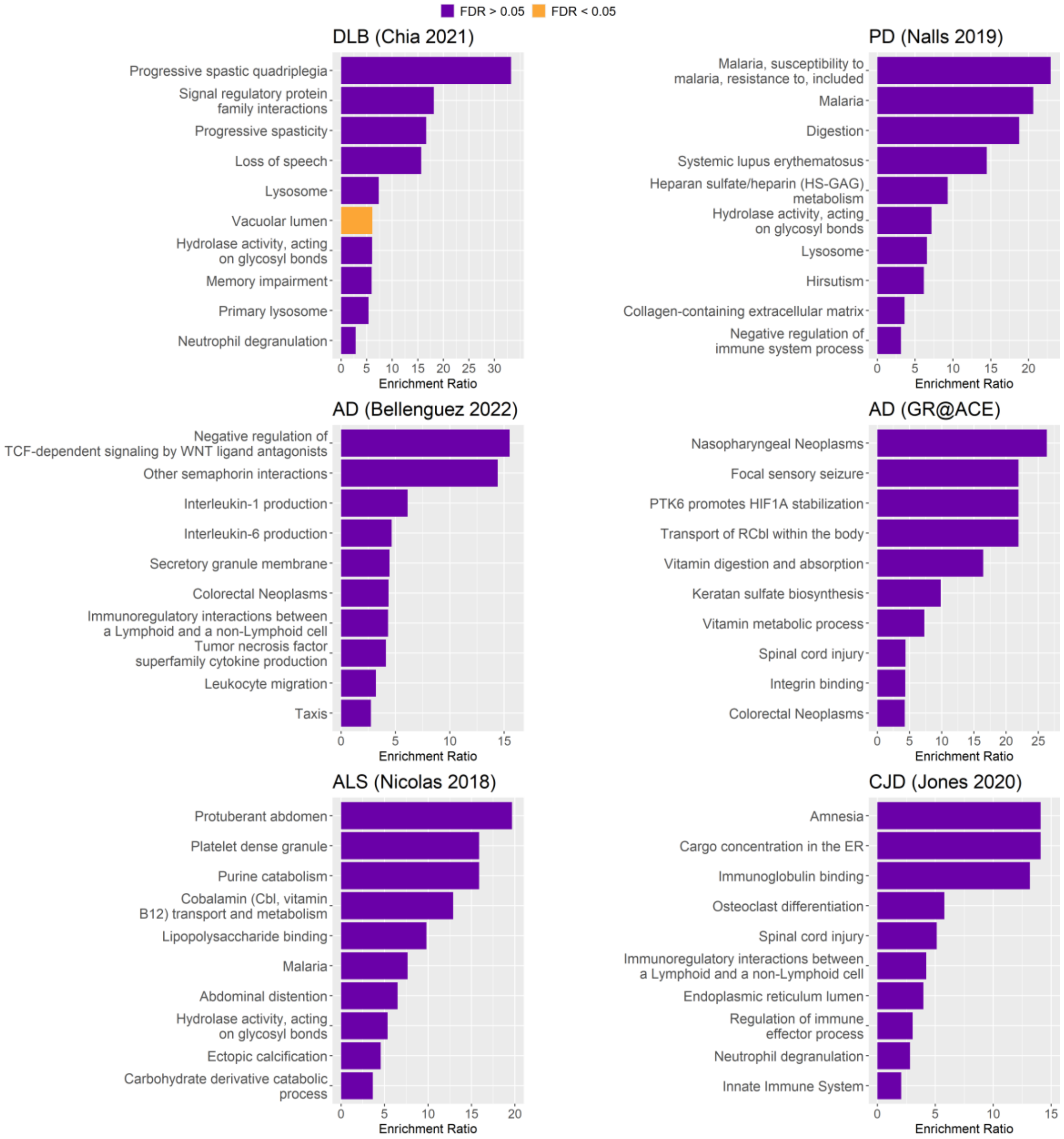


***Supplementary Figure 11. Top 10 enrichment analysis (ORA) of the top 100 ranking of reproducible proteins in MR results of neurodegenerative diseases.*** *The significance threshold was set to FDR<0.05 (orange).*

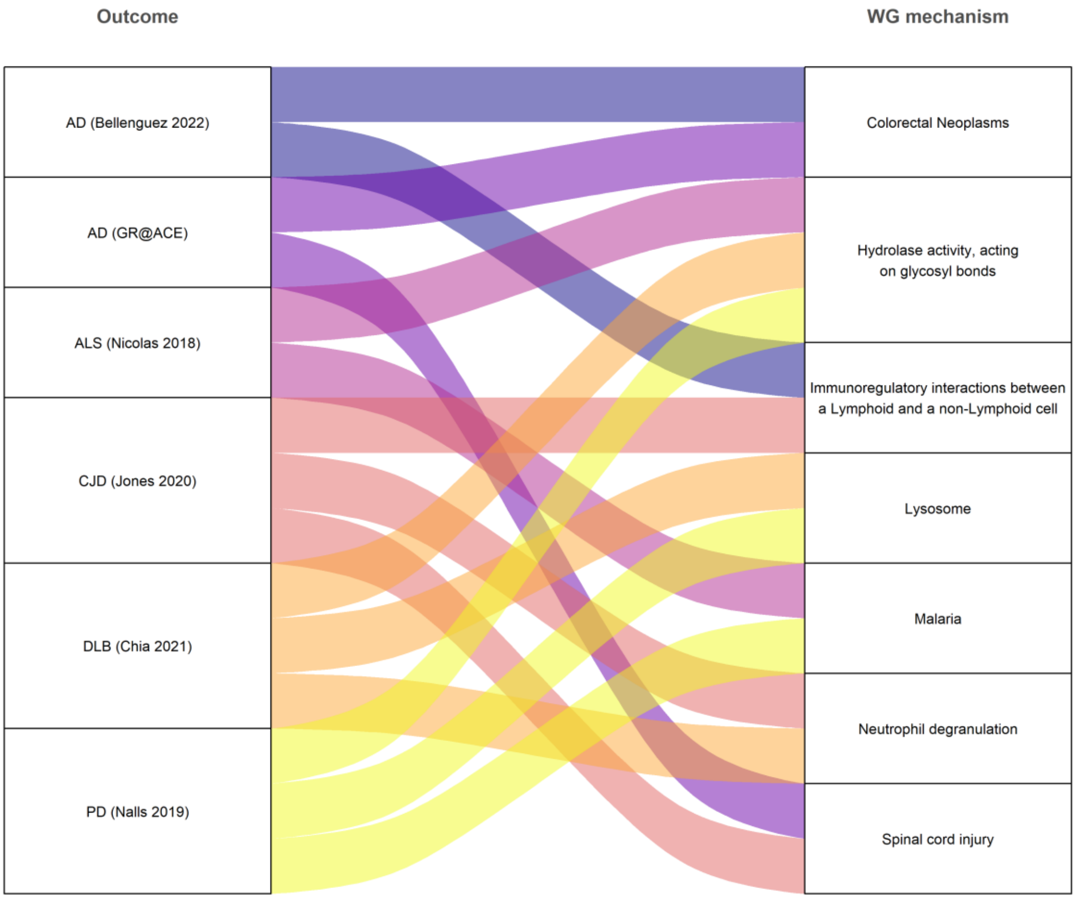


***Supplementary Figure 12. Overlapping mechanisms across neurodegenerative diseases considering only reliable proteins in the top 100 rankings.***

Considering all proteins

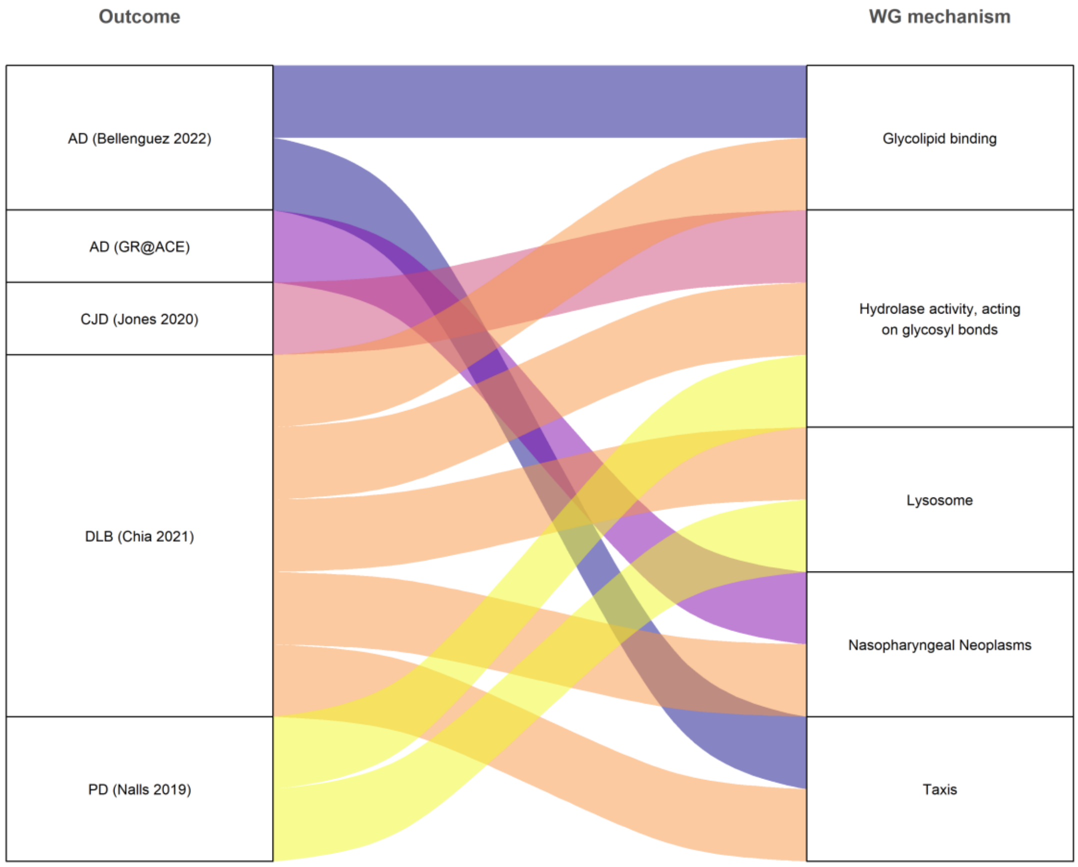


***Supplementary Figure 13. Enriched mechanisms represented across multiple diseases using the WebGestalt tool (WG).*** *These results were obtained considering the complete set of proteins with non-pleiotropic cis-pQTLs.*

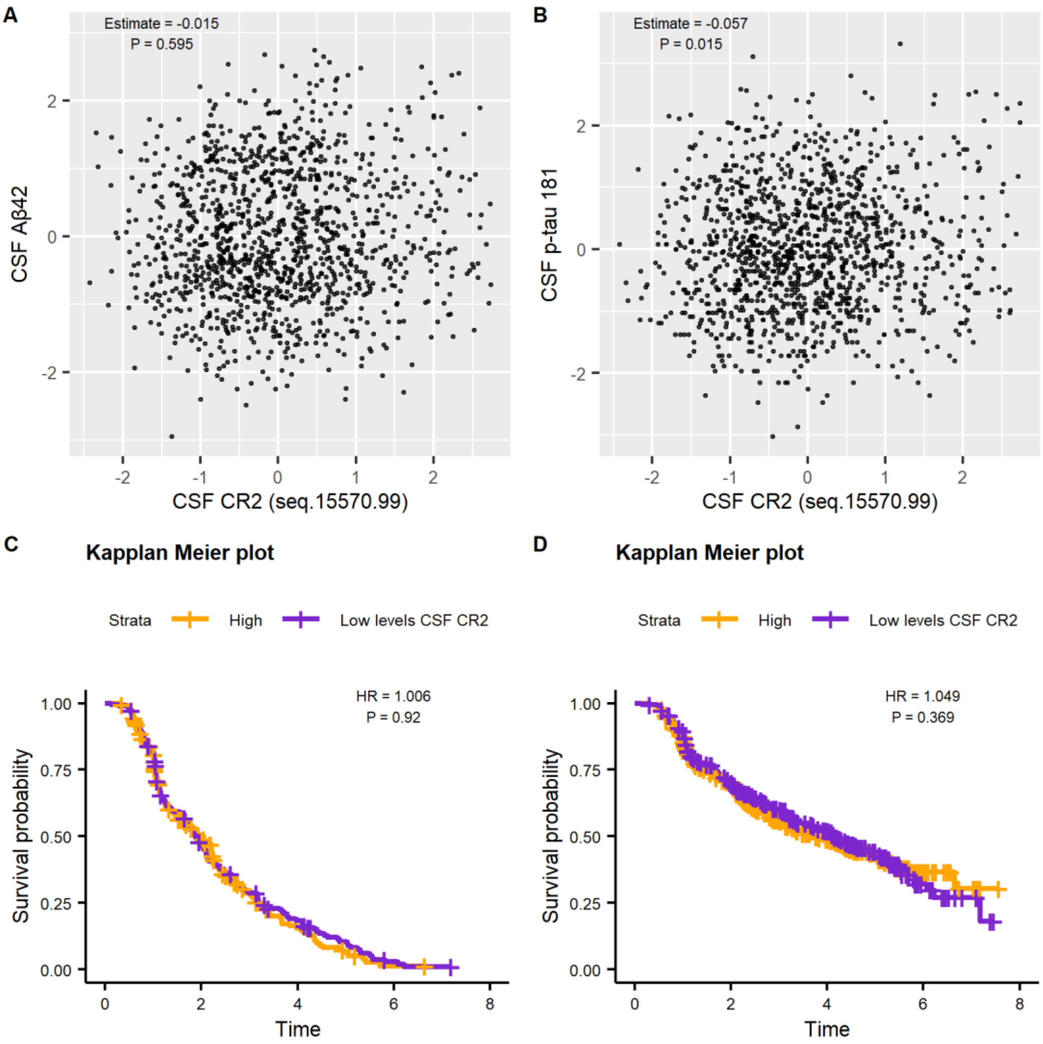


***Supplementary Figure 14. Characterization CSF CR2 levels.*** *A) Association between CSF CR2 and CSF Aβ42 levels (log-transformed and scaled). B) Association between CSF CR2 and CSF p-tau 181 levels (log-transformed and scaled). Estimates and P-values are reported for linear model adjusting by age, sex, the first 2 principal components. C) Cox proportional hazards regression model of the association between CSF CR2 levels and conversion to AD dementia. D) Cox proportional hazards regression model of the association between CSF CR2 levels and conversion to general dementia.*

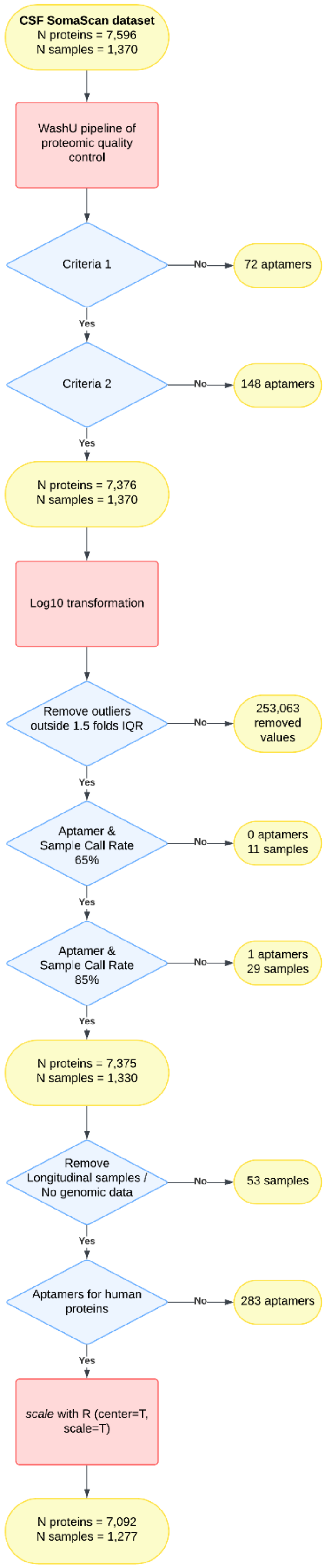


***Supplementary Figure 15. Flowchart of the proteomic quality control based on Western et al., 2024.***

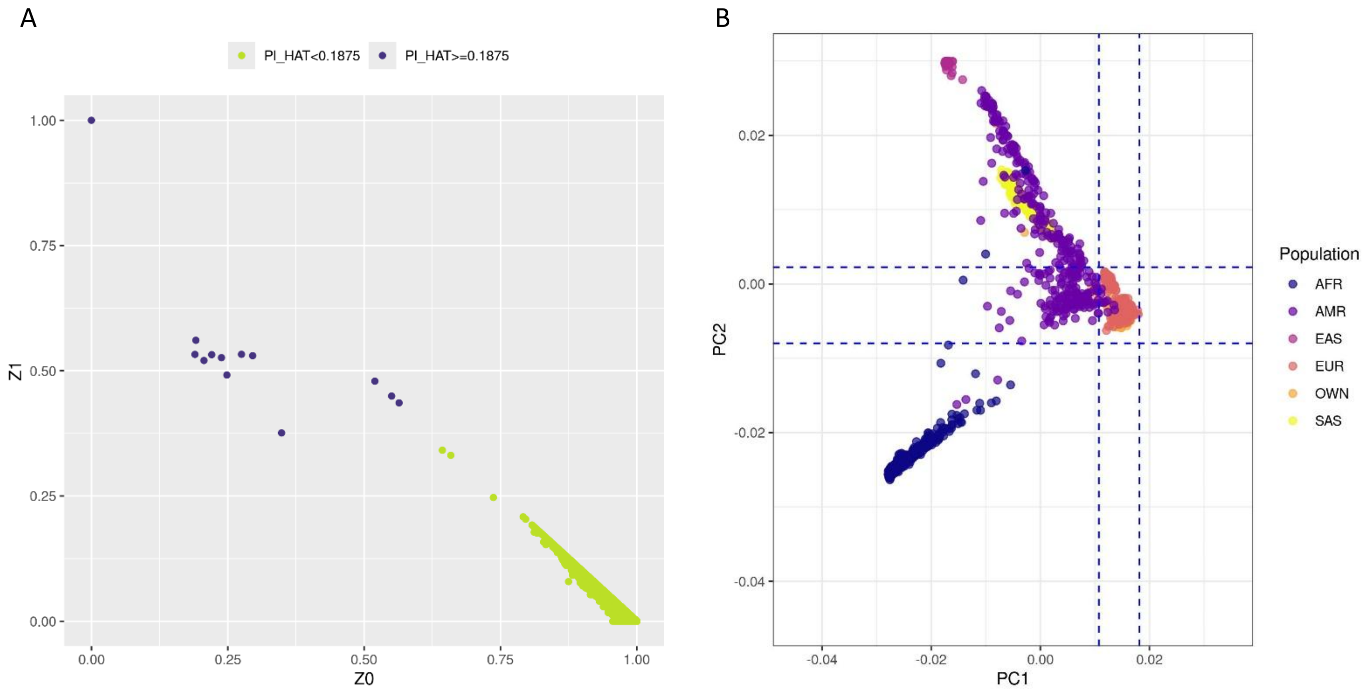


**Supplementary Figure 16. Genomic quality control and data processing.** A) Representation of the PI_HAT parameters from the IBD analysis. B) Representation of the PCA including 1000Genome data coloured by ancestry. The dashed lines represent the ±3 SD from the mean of individuals of European ancestry.

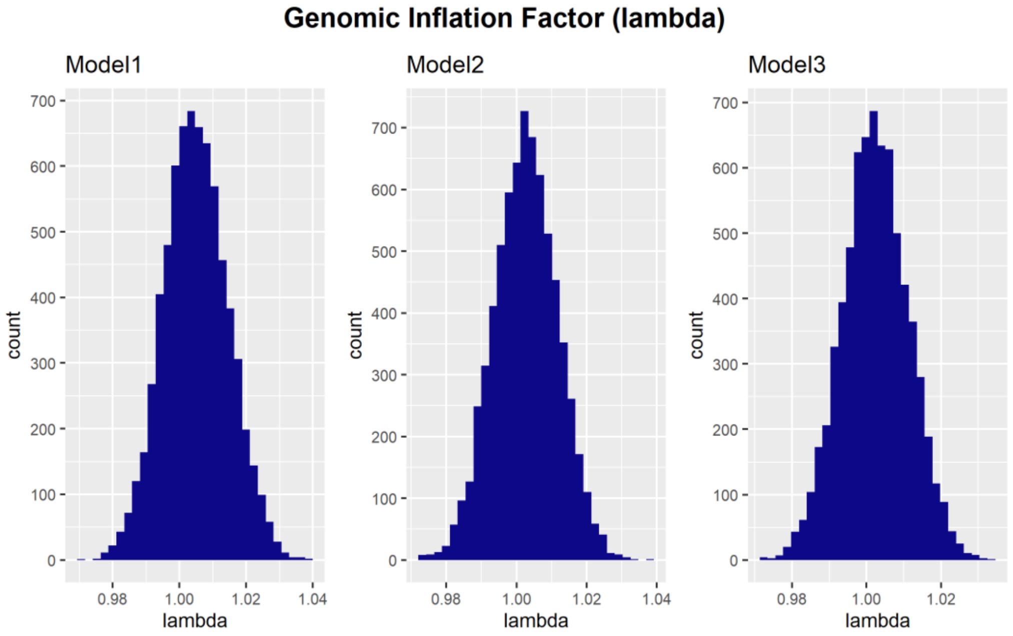


***Supplementary Figure 17. Genomic inflation factor (lambda) for the GWAS of the 3 models.***
